# A rapid review of the effectiveness of remote consultations versus face-to-face consultations in secondary care surgical outpatient settings

**DOI:** 10.1101/2022.09.04.22279537

**Authors:** Deborah Edwards, Judit Csontos, Liz Gillen, Judith Carrier, Ruth Lewis, Alison Cooper, Adrian Edwards

## Abstract

The use of remote consultations and telemedicine approaches significantly increased over the pandemic. There is evidence that some patients still prefer this mode of care delivery and time saving may also enable additional consultations and help to reduce waiting lists. However, the effectiveness of remote consulting for certain specialities, such as surgery, is unclear.

The aim of this review was to investigate the effectiveness of video or telephone consultations, particularly focusing on clinical, patient reported and safety outcomes, in adult secondary surgical outpatient care during the COVID-19 pandemic.

14 studies were identified. These were published in 2021-2022. Evidence is low or very-low quality due to observational study designs, small sample sizes and patient selection.

Policy and practice implications: Evidence is of low quality but suggests that for many surgical outpatient consultations, remote consultations are as effective as in-person consultations. There is potential for time and cost savings for remote consultations compared to in-person consultations. High quality research is needed to evaluate the effectiveness of remote consultations to understand which patients and which surgical specialities would benefit most.

**Funding statement:** The Wales Centre for Evidence Based Care was funded for this work by the Wales COVID-19 Evidence Centre, itself funded by Health & Care Research Wales on behalf of Welsh Government.

## 1. BACKGROUND

This Rapid Review is being conducted as part of the Wales COVID-19 Evidence Centre Work Programme. Question suggested by the All Wales Medical Directors, Aneurin Bevan Health Board and Royal College of Podiatry and refined to the above research question following stakeholder consultation.

### 1.1 Purpose of this review

Waiting times for elective treatments have increased since the emergence of COVID-19, as non-emergency treatments were suspended or delayed to focus on the pandemic response, creating a significant backlog for the National Health Service (NHS). As of February 2022, 6.18 million people were waiting for consultant led elective treatments to start in England, out of which 299,478 people have been on a waiting list for over a year, and 23,281 for over two years (Nuffield Trust 2022). In Wales, treatment waiting times follow a similar tendency, with 252,000 people waiting more than 36 weeks for treatment from referral (Welsh Government 2022).

To clear the backlog and reduce waiting times, new and innovative approaches are needed. There is some evidence from orthopaedic specialties and fracture clinics that telemedicine can free up time for healthcare professionals and enable medical staff to see more people, reducing waiting times (Jenkins & Halai 2021, Moisan et al. 2021). However, confidence in these findings is low or very low based on the quality of the studies (Jenkins & Halai 2021), thus more research might be required to see whether telemedicine can help decrease waiting times.

Moreover, uptake for telemedicine might be an issue, as in 2019, prior to the COVID-19 pandemic around 3.5% of outpatient appointments were provided via video or telephone UK-wide (Hutchings 2020). While at the start of the pandemic, the use of telemedicine approaches significantly increased (Gachabayov et al. 2022), with 35% of outpatient appointments conducted remotely in April 2020 UK-wide, following the easing of some pandemic restrictions, this percentage reduced to 25% in September 2020 (QualityWatch 2020). This indicates that in-person consultations might still be the preferred mode of healthcare provision. However, evidence suggests that some patients prefer telemedicine as the main source of care delivery (Technology Enabled Care Cymru 2021). Based on these changes in telemedicine provision brought on by the pandemic, the Welsh Government (2022a) has proposed an ambitious plan that aims for 35% of initial assessments, and 50% of follow-up appointments to be provided remotely. However, how telemedicine impacts the care given to patients, and the movement and management of patients through the outpatient cycle, particularly in a wide variety of surgical specialties is unclear.

Based on preliminary searches conducted for a rapid evidence summary, we found an umbrella review (Smith et al. 2021) and numerous systematic reviews investigating the use of telemedicine in surgical specialties as well as patient and/or provider satisfaction (Cabrera et al. 2020, Fahey et al. 2021, Gupta et al. 2021, Kolcun et al. 2020, Chaudhry et al. 2021, McMaster et al. 2021). However, in some of these systematic reviews the use of synchronous (video and telephone consultations) and asynchronous modalities (texting, e-mails, mobile applications, etc.) were often considered together, and separating their impact on outcomes was not possible (Smith et al. 2021). The umbrella review found that the most commonly reported outcome was patient satisfaction although this measure is not necessarily indicative of whether remote consultations and telemedicine was effective in improving clinical and patient reported outcomes (Smith et al. 2021). In addition, systematic reviews often included mixed populations of both adults and children, even though remote consultations for children can require a different approach, such as parents with young children having to describe symptoms in absence of physical examination (Tully et al. 2021). We also found missing or limited reporting of research conducted since the emergence of COVID-19 which would be particularly important as telemedicine use increased significantly during the pandemic. Across these reviews there is a plethora of information on usage, patient satisfaction but very little that has explored surgical outcomes. Therefore, this rapid review aims to investigate the effectiveness of remote consultations (video or telephone), particularly focusing on clinical, patient reported and safety outcomes, in adult secondary surgical outpatient care during the COVID-19 pandemic.

## 2. RESULTS

Of the 13,302 citations retrieved from our searches, **three prospective cohort studies and 11 retrospective cohort studies** met our eligibility criteria. The evidence is reported separately for the prospective studies and retrospective cohort studies. Further details about the prospective cohort studies are presented in Tables 1 (summary) and 2 (full data extraction).

**Table 1:**
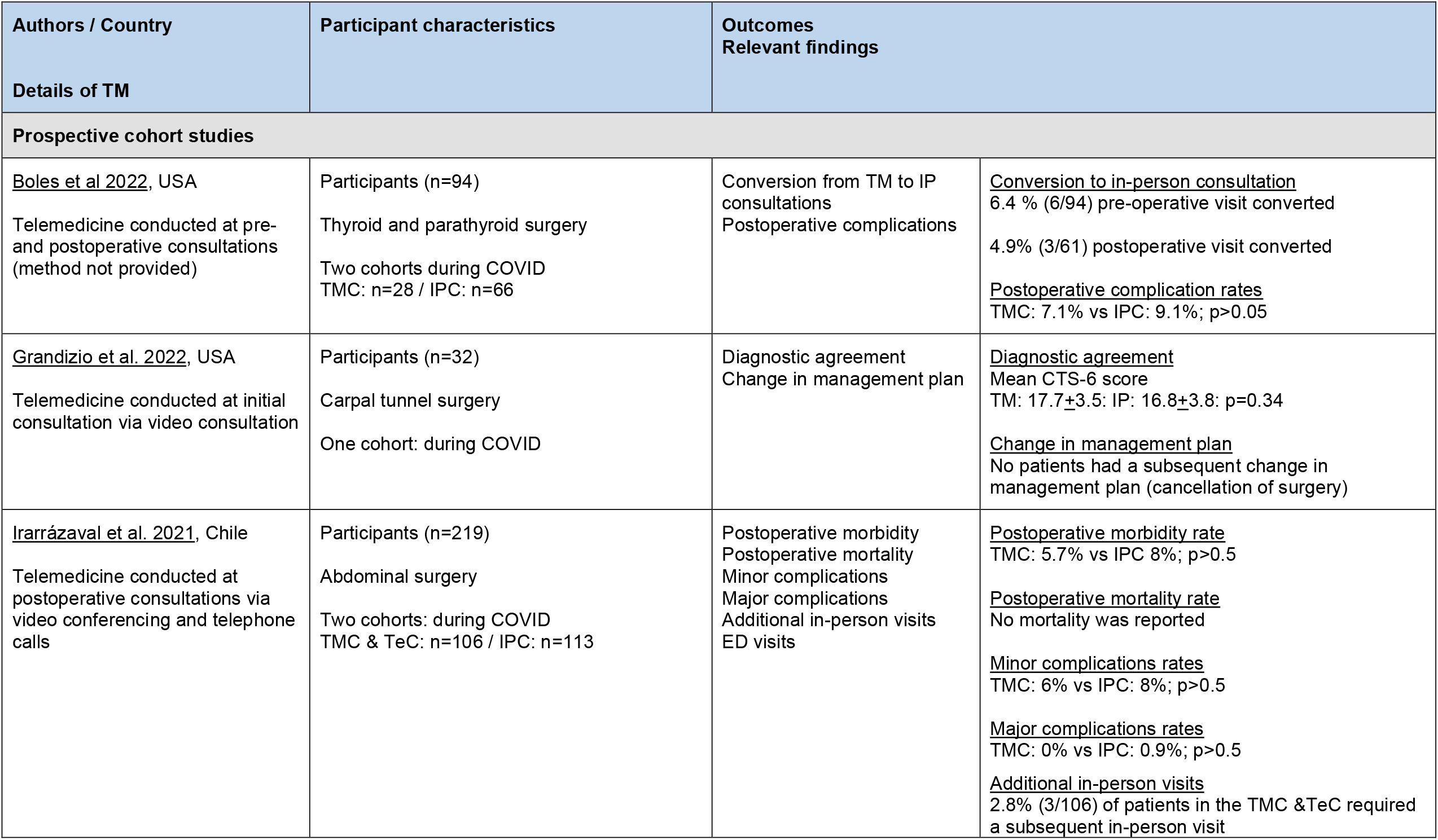
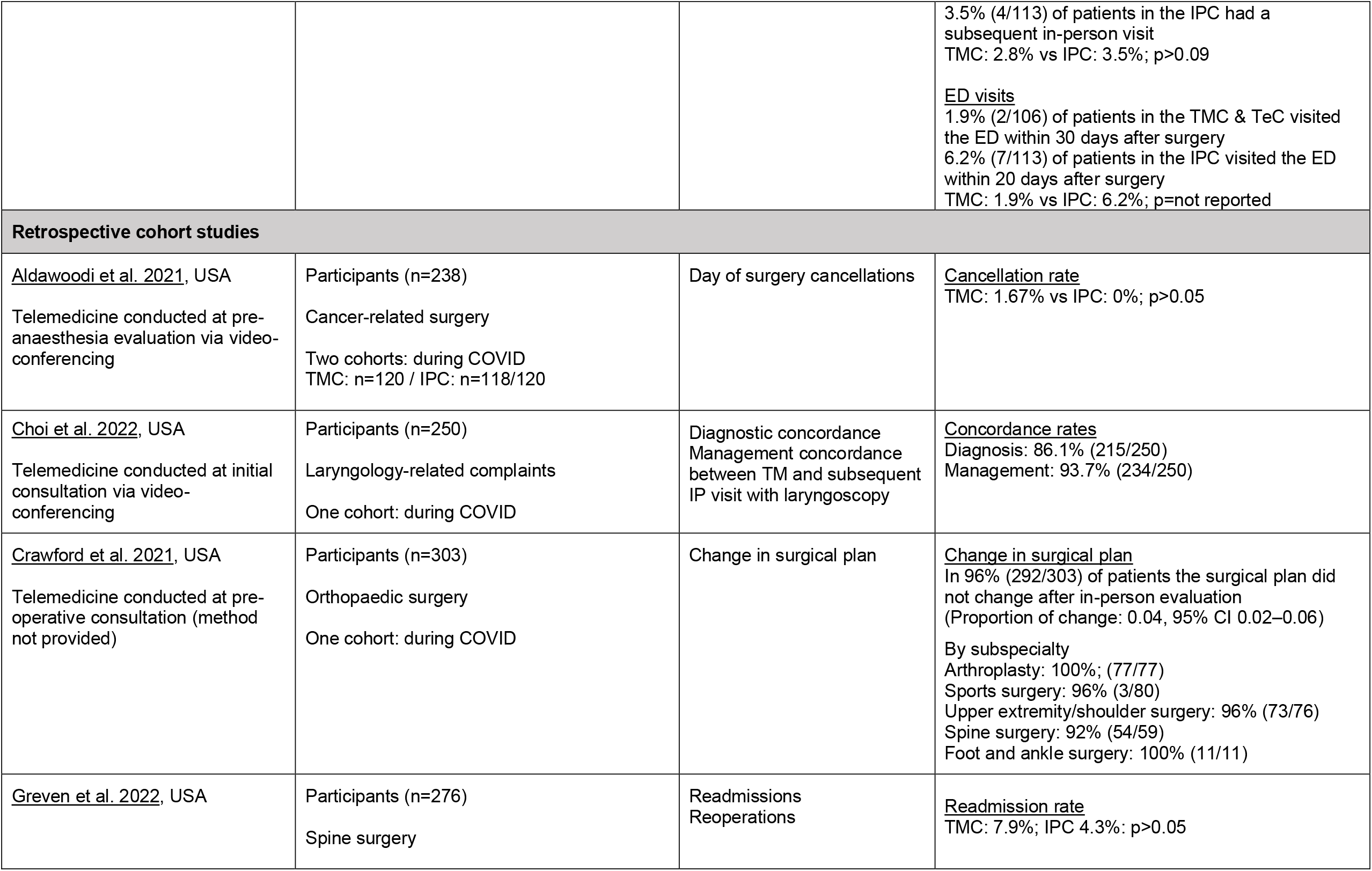
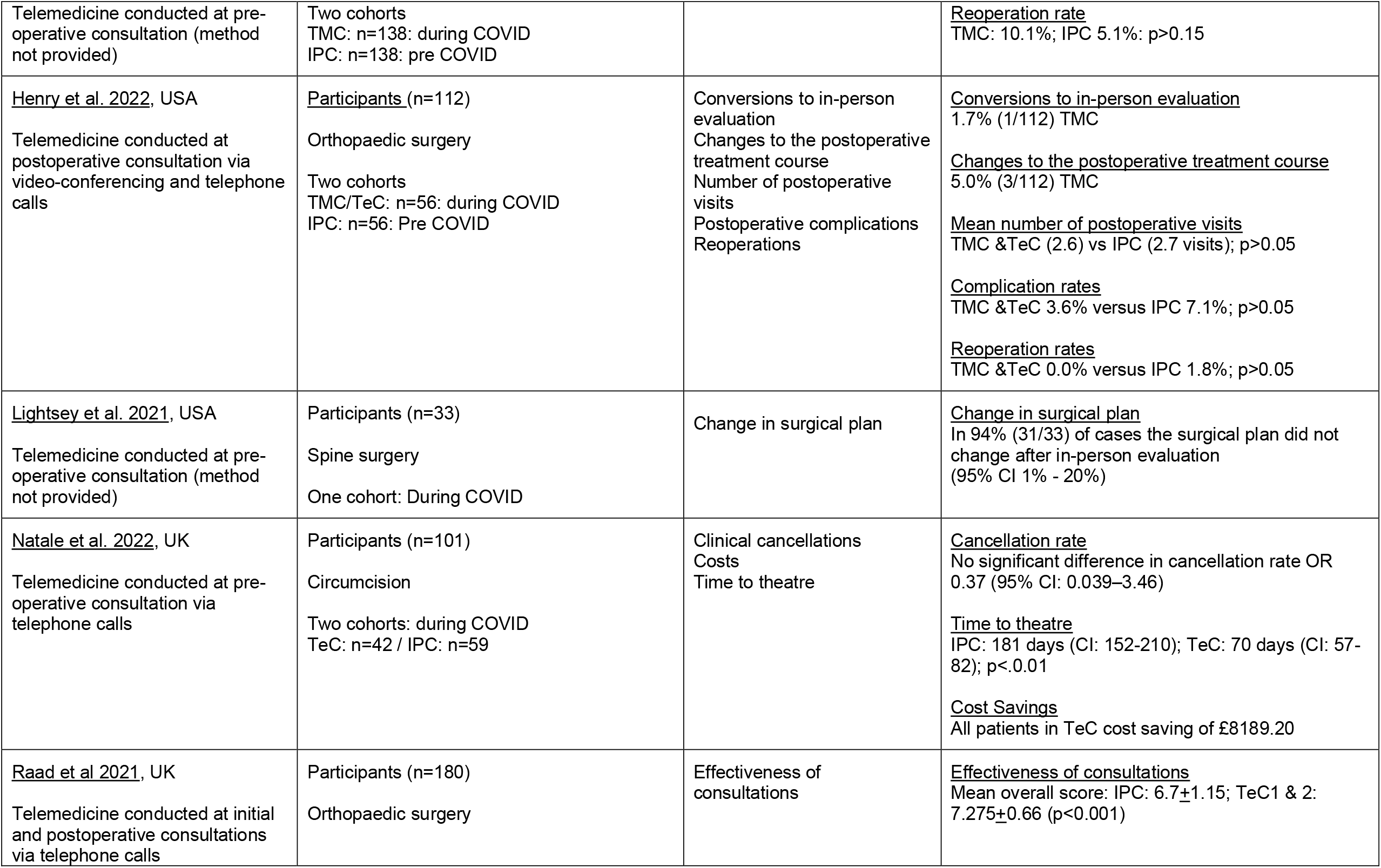
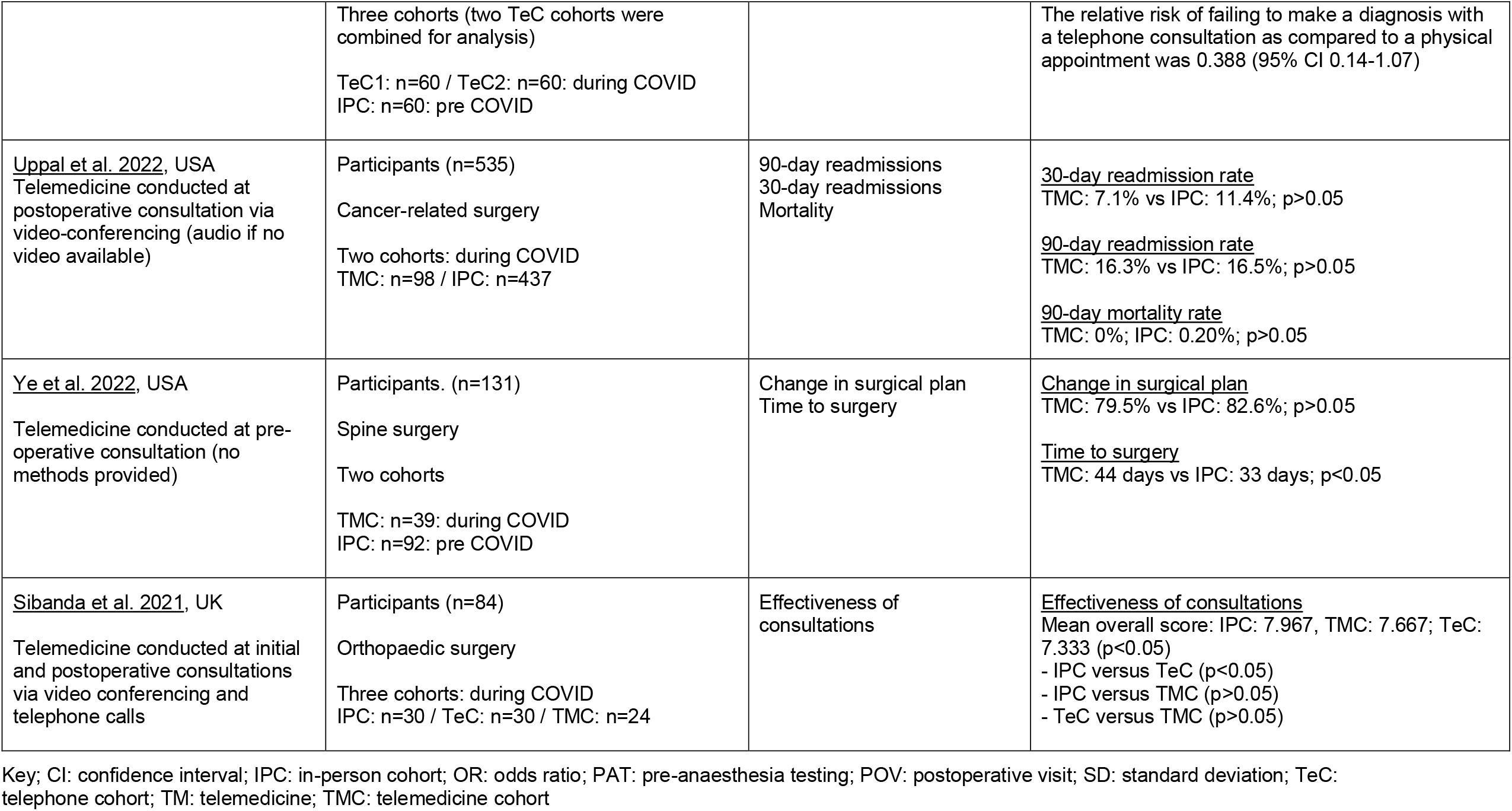
Summary of included studies.

### 2.1 Overview of the Evidence Base

#### 2.1.1. Prospective cohort studies

Two studies were conducted in the USA (Boles et al. 2022, Grandizio et al. 2022) and one in Chile (Irarrázaval et al. 2021). Numbers of participants across the studies ranged from 32 (Grandizio et al. 2022) to 219 (Irarrázaval et al. 2021). The studies were conducted with patients who were undergoing thyroid and parathyroid surgery (Boles et al. 2022), carpal tunnel release surgery (Grandizio et al. 2022) and abdominal surgery (Irarrázaval et al. 2021).

Eligibility for telemedicine was based on patients’ preference in two studies (Irarrázaval et al. 2021, Grandizio et al. 2022). In the third study, while patients could opt to have either telemedicine or in-person evaluation, the surgeons could decide to convert participants to in-person evaluation based on their clinical judgement and the patients’ medical history (Boles et al. 2022).

Telemedicine was used for initial consultations (Grandizio et al. 2022), postoperative consultations (Irarrázaval et al. 2021), and for both pre- and postoperative consultations (Boles et al. 2022) via video conferencing (Grandizio et al. 2022) and video conferencing or telephone calls (Irarrázaval et al. 2021). In one study however, no details were provided on the mode of telemedicine used (Boles et al. 2022).

The number of cohorts in the prospective studies differed. In one study, one cohort was used and the patient’s initial telemedicine consultation was compared to a second in-person consultation just before surgery (Grandizio et al. 2022). In two studies, two cohort groups (telemedicine and in-person) were compared (Boles et al. 2022, Irarrázaval et al. 2021).

The outcomes varied across the studies and included conversion from telemedicine to in-person consultation (Boles et al. 2022), postoperative complications (Boles et al. 2022, Irarrázaval et al. 2021), diagnostic agreement (Grandizio et al. 2022), change in management plan (Grandizio et al. 2022), additional in-person visits (Irarrázaval et al. 2021), emergency department visits within 30 days of surgery (Irarrázaval et al. 2021) postoperative morbidity (Irarrázaval et al. 2021) and mortality (Irarrázaval et al. 2021).

#### 2.1.2. Retrospective cohort studies

Eight studies were conducted in the USA and three in the UK (Natale et al. 2022, Raad et al. 2021, Sibanda et al. 2021). Numbers of participants across the studies ranged from 33 (Lightsey et al. 2021) to 535 (Uppal et al. 2022). Four studies were conducted with patients who were undergoing a variety of orthopaedic surgical procedures (Crawford et al. 2021, Henry et al. 2022, Raad et al. 2021, Sibanda et al. 2021), three studies for spine surgery (Lightsey et al. 2021, Greven et al. 2022, Ye et al. 2022), two studies for cancer-related surgery (Aldawoodi et al. 2021, Uppal et al. 2022) and one study each for circumcision (Natale et al. 2022) or laryngology-related complaints (Choi et al. 2022).

Eligibility for telemedicine in the included retrospective cohort studies was either based on patients’ preference (Choi et al. 2022), the surgeons’ decision (Henry et al. 2022), a patient-surgeon cooperative decision (Uppal et al. 2022) or an objective decision tool (Aldawoodi et al. 2021). In two studies, reasons for decision against telemedicine was mentioned, which excluded patients who were complex, high-risk, or needed emergency surgery (Lightsey et al. 2021, Greven et al. 2022). For five of the included studies, no details on eligibility for telemedicine was reported (Crawford et al. 2021, Natale et al. 2022, Raad et al. 2021, Sibanda et al. 2021, Ye et al. 2022).

Five studies reported on the use of telemedicine consultations for pre-operative appointments (Crawford et al. 2021, Lightsey et al. 2021, Greven et al. 2022, Ye et al. 2022, Natale et al. 2022), two studies for postoperative appointments (Uppal et al. 2022, Henry et al. 2022), two studies across initial and postoperative appointments (Raad et al. 2021, Sibanda et al. 2021) and one study for pre-anaesthesia evaluations (Aldawoodi et al. 2021). Three studies conducted the consultation via both video conferencing and telephone calls (Uppal et al. 2022, Henry et al. 2022, Sibanda et al. 2021), two studies via video conferencing (Choi et al. 2022, Aldawoodi et al. 2021) and two studies via telephone calls (Natale et al. 2022, Raad et al. 2021). In four studies no detail was provided regarding how telemedicine was conducted or what platform was used (Crawford et al. 2021, Lightsey et al. 2021, Greven et al. 2022, Ye et al. 2022).

The number of cohorts in the retrospective cohort studies differed. In three studies, one cohort was used (Choi et al. 2022, Crawford et al. 2021, Lightsey et al. 2021), and the patients’ initial telemedicine consultation was compared to a second in-person consultation. In seven studies, two cohort groups (telemedicine and in-person) were compared (Aldawoodi et al. 2021, Natale et al. 2022, Raad et al. 2021, Ye et al. 2022, Lightsey et al. 2021, Uppal et al. 2022, Greven et al. 2022, Henry et al. 2022) and in one additional study, three cohorts were compared (Sibanda et al. 2021). Comparison groups either contained participants who were chosen from a sample receiving in-person consultation during COVID or pre COVID historical patient pools.

The outcomes varied across the included studies with change in surgical plan being explored across three studies (Crawford et al. 2021, Lightsey et al. 2021, Ye et al. 2022). Time to surgery operation (Natale et al. 2021; Ye et al, 2022), day of surgery cancellations (Aldawoodi et al. 2021, Natale et al. 2022), consultation effectiveness (Raad et al. 2021, Sibanda et al. 2021), reoperations (Greven et al. 2022, Henry et al. 2022) were all reported across two studies. A number of other outcomes were explored which included conversions to in-person consultation (Henry et al. 2022), readmissions (Natale et al. 2022, Greven et al. 2022), and costs (Natale et al. 2022), mortality (Natale et al. 2022), postoperative complications (Henry et al. 2022), number of postoperative visits (Henry et al. 2022), changes to the postoperative treatment course (Henry et al. 2022) and diagnostic concordance between in-person and telemedicine visit (Choi et al. 2022).

### 2.2 Effectiveness of Telemedicine

To explore whether telemedicine consultations were as effective as in-person appointments in surgical specialties, results from the included studies are summarised below, with findings from prospective and retrospective studies presented separately by surgical subspeciality.

#### Prospective cohort studies

##### Thyroid and parathyroid surgery

A prospective cohort study of patients undergoing thyroid and parathyroid surgery at a tertiary care centre in a COVID-19 hotspot from March 2020 to October 2020 was undertaken by Boles et al (2022). The objective was to compare the safety and efficacy of telemedicine (n=28) with in-person pre-operative visits (n=66) in patients undergoing thyroid and parathyroid surgery. Patients were all offered an initial telemedicine consultation; however, they were also given the choice to request an in-person appointment. In addition, based on clinical presentation (large tumours, advanced cancer, voice or swallow change) some telemedicine patients were specifically selected for in-person evaluation by their surgeon. The outcomes of interest to this review were conversions to in-person consultations at pre- or postoperative consultations and postoperative complication rates. Six patients (6.4%) had their pre-operative visit converted from telemedicine to in-person and three (4.9%) had their postoperative visit converted from telemedicine to in-person due to a variety of clinical reasons or patient preference. There were no significant differences between the two cohorts in terms of postoperative complications (TMC: 7.1% vs IPC: 9.1%; p>0.05).

##### Gastrointestinal surgery

Telemedicine clinics compared to in-person follow up for postoperative care after gastrointestinal surgery during the Covid-19 outbreak were the focus of the prospective study conducted by Irarrázaval et al. (2021). Patients were given the option of telemedicine follow up, 106 (48%) opted for telemedicine and 113 (52%) for an in-person consult. There were no significant differences in postoperative morbidity rate (TMC: 5.7% vs IPC 8%; p>0.50); minor complications rate (TMC: 6% vs IPC: 8%; p>0.05) or major complications rate (TMC: 0% vs IPC: 0.9%; p>0.05). No postoperative mortality was reported for either group. Additionally, there were no significant differences in additional postoperative in-person visits (TMC: 2.8% vs IPC: 3.5%; p>>0.05), or visits to the emergency department within 30 days of surgery (TMC: 1.9% vs IPC: 6.2%; p=not reported).

##### Carpal tunnel syndrome

A prospective evaluation of telemedicine for routine referral for carpal tunnel syndrome (CTS) pre-surgery was conducted by Grandizio et al (2022). Patients referred for CTS were offered a telemedicine pathway and 32 were included. A modified CTS-6 instrument as part of telemedicine screening for patients being evaluated for CTS was compared to in-person administration of the conventional CTS-6 instrument with the same group of patients. The conventional CTS-6 is a six-item instrument for the diagnosis of CTS. CTS-6 components focus on the examination of numbness in the median nerve distribution, nocturnal numbness, thenar atrophy or muscle weakness, Phalen’s test, two-point discrimination, and the Tinel sign. During an in-person consultation, the surgeon would perform these tests, thus CTS-6 was modified that elements can be performed by patients at a telemedicine visit. After a diagnosis and management plan was developed during the telemedicine visit, study participants indicated for surgery were also evaluated in-person on the day of surgery. There were no significant differences in mean CTS-6 scores completed during the telemedicine and in-person consultations. There was diagnostic agreement across the different tests and manoeuvres performed, although telemedicine examination of median nerve sensory changes demonstrated lower levels of agreement than in-person evaluation. There were no subsequent changes in management plan (cancellation of surgery) based on in-person evaluation.

#### Bottom line results from prospective cohort studies

This section summarised the effectiveness of telemedicine derived from three prospective cohort studies.

Only small numbers of patients undergoing **thyroid and parathyroid surgery** had their pre- or **postoperative consultations converted** from telemedicine to in-person. Very low quality evidence from two prospective cohort studies suggests that the presentation of **postoperative complications** at the postoperative consultation are **similar** regardless of the mode of consultations (telemedicine compared to in-person) for **thyroid and parathyroid or abdominal surgery**.

Very low quality evidence from one prospective cohort study of patients undergoing **abdominal surgery** suggests that the presentation of **postoperative morbidity** at the postoperative consultation, visiting the **ED within 30 days** of surgery or the need for an **additional in-person visits** are **similar** regardless of the mode of consultation (telemedicine compared to in-person). Additionally, **no postoperative mortality** was reported for either group.

Additionally, low quality evidence from one prospective study shows that there was **diagnostic agreement** across the different tests and manoeuvres performed during initial telemedicine consultations and subsequent in-person examinations of **carpal tunnel syndrome**, although median nerve sensory change tests might need further adjusting to telemedicine administration. None of the patients evaluated for **carpal tunnel syndrome** during an initial telemedicine consultation had a **change in management plan** (cancellation of surgery) after a subsequent in-person consultation.

#### Retrospective cohort studies

##### Orthopaedic surgery

Two studies (Raad et al. 2021, Sibanda et al. 2021) used the Ashford Clinic Letter Scoring system (Virani et al. 2021), a tool validated to assess remote clinic appointments by research or medical staff, to measure the quality and efficacy of consultations within the same orthopaedic setting. This system rates four consultation parameters (making diagnosis, investigations, formulation of a treatment plan, and value of consultation) from zero to two. With each parameter scored, a maximum total score of eight can be achieved, which indicates a highly effective consultation. It was reported that there were no differences in overall scores between those who had their initial or follow-up appointments (data combined) by video conferencing compared to in-person (IPC: 7.967; TMC: 7.667; p>0.05) (Sibanda et al. 2021). However, across the two studies conducted with different participants through different timeframes there were conflicting results with Sibanda et al. (2021) reporting that patients seen via telephone consultations during COVID scored significantly lower than those seen in-person during COVID (IPC: 7.967; TeC: 7.333; p<0.05). Whereas Raad et al. (2021) reported that patients seen via telephone consultations during COVID scored significantly higher than those seen in-person pre-COVID (IPC: 6.7; TeC: 7.275; p<0.001).

Crawford et al. (2022) reported a 96% (292/303) surgical plan accuracy between the initial telemedicine visit and an in-person visit. Plan changes (11/303) were attributed to patient preference, additional imaging or adding components to the surgical plan bases on surgeon’s recall. Plan changes were reported for patients scheduled for sports (3.75%; 3/80), upper extremity/shoulder (3.95%; 3/76), spine (8.47%; 5/59) surgery with were no plan changes for patients scheduled for joint arthroplasty (0.00%; 0/77) and foot and ankle surgery (0.00%; 0/11). There was notable variability in the conduct of virtual examinations across subspecialties.

One study (Henry et al. 2022) reported on whether change to in-person consultation was necessary after the initial postoperative telemedicine consultation (video conferencing or telephone call) following upper extremity surgery. Change to in-person evaluation was to further address a specific complaint or concern due to the intrinsic limitations of virtual consultations, such as inadequate physical examination. One (1.7%) telemedicine consultation required conversion to an in-person evaluation, this was due to suspected superficial infection necessitating an in-depth physical examination. The postoperative treatment course required a specific change solely based on the findings of the telemedicine visit for 5.0% (3/112) of cases. Postoperative complication rates (TMC & TeC: 3.6%; IPC: 7.1%); postoperative visits (TMC & TeC: 2.6 visits; IPC: 2.7 visits); and reoperation rates (TMC & TeC: 0.0%; IPC: 1.8%) were also reported, and no significant differences were found between the virtual and in-person cohorts (p>0.05).

##### Spine surgery

Two studies explored whether telemedicine and in-person evaluations generated similarly accurate surgical plans and whether these plans were subject to change (Lightsey et al. 2021, Ye et al. 2022). Change was defined as a change in the extent of surgery offered, in the approach, in type of surgery or region of surgery (Lightsey et al. 2021, Ye et al. 2022) and whether the patient previously indicated for surgery was found not to merit a surgical procedure (Lightsey et al. 2021). Lightsey et al. (2021) found that for 94% of cases the surgical plan did not change after subsequent in-person evaluation (95% CI 1% - 20%) and Ye et al. (2022) reported that telemedicine (79.5%) and in-person (82.6%) evaluations generated similarly accurate surgical plans that did not need to change on the day of surgery (p>0.05). However, in Ye et al.’s study (2022) the telemedicine cohort experienced significantly longer time between the initial appointment and surgery (TMC: 44 days; IPC: 33 days; p<0.05) compared with the in-person cohort. Greven et al. (2022) found no significant differences in readmission (TMC: 7.9%; IPC: 4.3%; p>0.05) or reoperations rates (TMC: 10.1%; IPC: 5.1%; p>0.05) for spine surgery candidates evaluated pre-operatively by telemedicine compared to in-person.

##### Cancer-related surgery

Aldawoodi et al. (2021) investigated how many surgical procedures needed to be cancelled when the pre-anaesthesia evaluation was conducted via video-conferencing. It was reported that there were no differences in the cancelation rates compared to an in-person cohort (TMC: 1.67%; IPC: 0%; p>0.05). When postoperative consultations were conducted via telemedicine, Uppal et al. (2022) reported that there were no differences in the 30-day readmission rate (TMC: 7.1%; IPC: 11.4%; p>0.05), the 90-day re-admission rate (TMC: 16.3%; IPC: 16.5%; p>0.05) or 90-day mortality rate (TMC: 0%; IPC: 0.20%; p>0.05).

##### Laryngology-related complaints

The concordance in diagnosis and management between initial consultations conducted via video-conferencing and the subsequent in-person visits with laryngoscopy for laryngology-related complaints was investigated by Choi et al. (2022). Concordance rates for diagnosis were 86.1% and for management 93.7%.

##### Circumcision

Natale et al. (2022) sought to determine whether pre-operative telephone calls are an effective alternative to in-person assessment of patients requiring circumcision. Cases cancelled from the operating lists and the rationale for cancellation were recorded and defined as any cancellation related to patient health, operative or anaesthetic factors. There were no significant differences in cancellation rate (OR 0.37; 95% CI: 0.039–3.46). Time to treatment was decreased in the telephone clinic group (IPC: 181 days; TeC: 70 days; p<0.01). Telephone clinic could have achieved a per-patient cost reduction of £81 and a total cost savings of around £8,200 if all participants had been assessed via telemedicine.

#### Bottom line results from retrospective cohort studies

This section summarised the effectiveness of telemedicine derived from 11 retrospective cohort studies.

Very low quality evidence from one retrospective study found that **day-of-surgery cancellation rates** were similar when **pre-anaesthesia evaluations** were conducted via **video-conferencing** compared to in-person for patients scheduled for **cancer-related surgery**. One further very low quality retrospective cohort study reported that postoperative **readmission and mortality within 30 and 90 days** following **cancer-related surgery** are **similar** regardless of the mode of consultation (telemedicine compared to in-person).

**Very low quality evidence from another retrospective cohort study showed that surgical plans** generated for **orthopaedic patients** are **rarely changed** by in-person evaluation. Very low quality evidence from one retrospective study suggests **video conferencing** was just as effective as in-person consultations for new referral and follow-up **orthopaedic patients** (surgical and non-surgical). Whereas for two additional retrospective studies there was **mixed** low quality evidence so the **effectiveness of telephone consultations** compared to in-person consultations **could not be determined**. Very low quality evidence from another retrospective cohort study reported that **postoperative complication rates, postoperative visits, reoperation rates** following **orthopaedic surgery** are **similar** regardless of the mode of consultation (telemedicine and telephone calls compared to in-person). Only small numbers of patients undergoing orthopaedic surgery required change to in-person consultation. Additionally, only a small number of patients required a specific change in the postoperative treatment course solely based on the findings of the telemedicine.

Very low quality evidence from one retrospective study found that **readmission and reoperations rates** were similar when **pre-operative consultations were** conducted via **video-conferencing** compared to in-person for patients scheduled for **spine surgery**. Additionally, very low quality evidence from one retrospective cohort study suggests that both video-conferencing and in-person evaluations **generated accurate spine surgical plans** that did not need to change on the day of surgery.

Very low quality evidence from one retrospective cohort study suggests that **costs** can be saved, and **time to surgery** decreases when **pre-operative consultations** are conducted via **telephone calls** compared to in-person for patients undergoing assessment for **circumcision**. Additionally, **clinical cancellation rates** were similar for both groups.

Very low quality evidence from one retrospective study suggests that telemedicine can be used to provide a **preliminary diagnosis and management plan** for **laryngology-related complaints**.

## 3. DISCUSSION

### 3.1 Summary of the findings

Previous reviews (Petersen et al. 2021, Gupta et al. 2021, Kolcun et al. 2020, Fahey et al. 2021, McMaster et al. 2021, Chaudhry et al. 2021) and a review of systematic reviews (Smith et al. 2021) have explored the usage, effectiveness or cost-effectiveness of telemedicine across a variety of surgical specialities prior to the COVID-19 pandemic. These have demonstrated the feasibility of the use of telemedicine for perioperative and/or postoperative care for adults and paediatric undergoing a variety of surgical procedures and overall, patients are satisfied with telemedicine in surgical practice. High patient and provider satisfaction for the use of telemedicine across a range of surgical specialities has previously been reported and is comparable to satisfaction obtained from in-person consultations (Smith et al. 2021, Fahey et al. 2021, McMaster et al. 2021, Chaudhry et al. 2021). There is a lack of evidence however, from existing systematic reviews of telemedicine for adult patients that have focused solely on the effectiveness of telemedicine during, and post COVID-19. A scoping review that mapped the evidence for telemedicine in surgical settings for the first year of the COVID-19 pandemic, reported a large increase in outpatient management (virtual clinics), new patient consultation, telesurgery use in education, followed by preoperative evaluation/triage (Gachabayov et al. 2022). However, in the first 6 months of the COVID-19 pandemic a lack of intervention studies was noted, and effectiveness was not reported. Therefore, this rapid review sought to investigate the effectiveness of remote consultations (video or telephone), particularly focusing on clinical, patient reported and safety outcomes, in adult secondary surgical outpatient care during the two years of COVID-19 pandemic.

The findings of this rapid review are based on very limited low and very low quality evidence (as determined using the GRADE approach) from three prospective and 11 retrospective cohort studies. However, a number of included studies made definitive claims about the effectiveness of the telemedicine consultations in surgical specialties. So, the findings from the included prospective and retrospective studies should be interpreted with caution.

The surgical specialities that are covered include thyroid and parathyroid gastrointestinal, orthopaedic, spine or cancer related surgery, circumcision, carpal tunnel release and surgery for laryngology-related complaints. Ye et al. (2022) comments that spinal surgeons may not be able to give an accurate diagnosis and formulate the correct surgical plan during a pre-operative telemedicine consultation. However, findings from two retrospective studies conducted with spine patients (Lightsey et al. 2021, Ye et al. 2022) and one further study with orthopaedic patients that included 59 spine patients (Crawford et al. 2021) all found that the plans generated during telemedicine consultations were as accurate as surgical plans generated at in-person consultations. Ye et al. (2022) comments that this is a surprising result given the importance of a detailed physical examination in spine practice. For laryngology related complaints pre-operative telemedicine can be used to provide a preliminary diagnosis and management plan and appropriate triaging. Surgical plans generated for orthopaedic patients, thyroid and parathyroid surgery are rarely changed by in-person evaluation and day-of-surgery cancellation are similar regardless of the mode of consultation showing telemedicine to be a feasible alternative to in-person consultations. Due to the limitations related to the study methodologies used there is a need for RCTs to be conducted that take into account confounding factors.

Pre-operative consultations have the potential to reduce the length of time a patient is waiting for surgery. Time to surgery was investigated across two studies in this rapid review (Natale et al, 2022; Ye et al. 2022) and presented mixed findings but this was due to the inclusion of a historical control group (pre-COVID) in one of the studies (Ye et al. 2022) so similar comparisons could not be made. Further studies with concurrent cohorts are needed to investigate this outcome further.

Only one study reported on costs, specifically on the cost savings associated with telephone consultations. So, no firm conclusions can be reached on the cost effectiveness of telemedicine consultations that were conducted during the COVID-19 pandemic.

Postoperative telemedicine consultations across a range of surgical procedures were found to be feasible with similar results reported across outcome measures compared to in-person consultations. However, across the included studies a patient’s eligibility for pre—operative or postoperative telemedicine was not always reported. Where this information was provided, telemedicine group allocation was either based on patients’ preference or surgeons’ clinical judgement. When clinical judgement for pre-operative patient selection was made then the patients who were deemed to be more complex or high-risk were excluded. There are also limitations across the studies where the patients could choose between in-person or telemedicine consult after surgery. This has the potential for bias, as patients who have easier access to technology or more confidence in using video or telephone applications might be favoured. Future studies should consider transparent reporting of telemedicine eligibility.

### 3.2 Limitations of the available evidence

The included studies had several limitations based on the methodological assessment. Two of the three prospective cohort studies (Boles et al. 2022, Irarrazaval et al. 2021) met seven criteria out of the 11 on the critical appraisal checklist. One further prospective cohort study scored six out of a potential nine criteria (Grandizio et al. 2022). Two questions were not applicable, as comparisons were not made between two populations (Q1, Q2), but between the same participant group receiving different interventions (telemedicine against face-to-face assessment) at different timepoints. In one study (Boles et al. 2022) the study groups in were not fully comparable at baseline (Q1) and for another study (Irarrazaval et al. 2021) it was unclear if the outcomes were measured in a valid and reliable way (Q7). None of the three prospective studies had sufficient strategies to deal with confounding factors (Q4) and had issues or a lack of information on whether follow-up of participants was complete (Q9) and strategies to deal with incomplete follow-up (Q10).

For the 11 retrospective cohort studies all the studies addressed an appropriate and clearly focused question (Q1.1), had clearly defined outcomes (Q1.7), reliable methods of assessment of exposure (Q1.10) and valid and reliable methods of outcome assessment (Q1.11). Out of six retrospective cohort studies, in which there were a comparison group, only one study (Henry et al. 2022) selected participant groups that were comparable in all aspects other than the factors under investigation (Q1.2). Only one retrospective cohort study (Choi et al. 2022) gave recognition that knowledge of exposure status could have influenced the assessment of outcome (Q1.9). While these are retrospective studies, and so neither the participants, nor the clinicians could have been blinded, methods to try and conceal group allocation from assessors could have been attempted, or bias arising from the assessors knowing allocation could have been disclosed. Only two studies (Natale et al 2022, Uppal et al. 2022) identified and accounted for potential confounders in the design and analysis (Q1.13). Fives studies provided confidence intervals (Q1.14) (Choi et al. 2022, Crawford et al. 2021, Lightsey et al. 2021, Natale et al. 2022, Uppal et al. 2022) and one further study provided confidence intervals for one of the outcomes but not for the others, so this had to be scored a ‘No’ (Raad et al. 2021). Overall, five of the retrospective cohort studies were rated as being of low quality reflecting that either most criteria were not met, or that there were significant flaws relating to key aspects of study design (Aldawoodi et al. 2021, Greven et al. 2022, Raad et al. 2021, Sibanda et al. 2021, Ye et al. 2022). This means that conclusions are likely to change in the light of further studies. A further six of the retrospective cohort studies were rated as being acceptable reflecting that most criteria were met and that there were some flaws in the study with an associated risk of bias (Choi et al. 2022, Crawford et al. 2021, Henry et al. 2022, Lightsey et al. 2021, Natale et al. 2022, Uppal et al. 2022). This means that conclusions may change in the light of further studies.

The sample sizes across the telemedicine arms of the included studies ranged from 32 (Grandizio et al. 2022) to 109 (Uppal et al. 2022). Only one study conducted a sample size calculation (Grandizio et al. 2022). In the absence of sample size calculations there is a need to be cautious when interpreting findings from studies. Additionally, strong conclusions should not be drawn due to the small sample sizes which are unlikely to produce reliable results.

### 3.3 Implications for policy and practice

Practitioners should ensure all new telemedicine initiatives undergo thorough service evaluation to ensure they meet the needs of both the speciality and the patients including patient reported outcomes.

Policy makers should ensure that any recommendations for telemedicine take into account the need for all types of service users.

Further high-quality research studies that take into account confounding factors should be funded to determine both clinical effectiveness and cost effectiveness

### 3.4 Strengths and limitations of this Rapid Review

The strength of this review is that a thorough search was undertaken by an information specialist across six electronic databases. Although this was a rapid review in which a number of the systematic review processes were streamlined, it should be noted that full-text screening, data extraction and critical appraisal of each study were undertaken by different reviewers but independently checked for accuracy and consistency by the same second reviewer, which is a strength of this work. Moreover, 20% of title and abstract screening conducted by one reviewer was checked by another reviewer to make sure that study selection was accurate and relevant records were identified.

Potential limitation of this rapid review is that even though, the accuracy of a portion of title and abstract screening was checked, it is possible that relevant records might have been missed in the group of records that was not double screened. In addition, due to time constraints of the rapid review process, extraction of some data had to be omitted, such as comorbidities. Therefore, it is possible that results are influenced by participants’ other health conditions. However, it must be noted that information on comorbidities were not available in all studies, thus extraction of this would not have been possible.

This rapid review was limited to studies published between 2020 and 2022, therefore it is possible that by including studies conducted before the COVID-19 pandemic might change the conclusions made in this report. However, this rapid review is reflective of the research conducted during the pandemic, and it could provide important insights into the use of telemedicine during a public health emergency. Moreover, this rapid review highlights gaps in the literature published since the start of the COVID-19 pandemic, which is a strength as it could help focus on areas where further high-quality research is needed.

Another limitation of this rapid review is the heterogeneity in the included studies, which is present in the different surgical specialties, various outcomes collected, and different video or telephone applications used for telemedicine. Therefore, pooling results to show whether telemedicine was effective was not possible. Furthermore, the methods to investigate the effectiveness of telemedicine consultations was varied. This variability could be noticed in the use of comparison groups, as while most of the included studies had two or more cohorts, a few only had one cohort and utilised within-subject research design. Moreover, many studies with two cohorts had pre-COVID in-person groups which makes direct comparison with studies using face-to-face controls recruited during COVID difficult.

## Data Availability

All data produced in the present study are available upon reasonable request to the authors

**Rapid Review Details**

**Review conducted by:**

Wales Centre for Evidence Based Care

**Review Team:**

- Deborah Edwards
- Judit Csontos
- Liz Gillen
- Judith Carrier

**Review submitted to the WCEC in:** July 2022

**Stakeholder consultation meeting:** 18^th^ July 2022

**Rapid Review report issued by the WCEC in:** August 2022

**WCEC Team:** Adrian Edwards, Alison Cooper, Ruth Lewis, Emma Small, Jane Greenwell and Micaela Gal, involved in drafting the Topline Summary and editing

**This review should be cited as:**

RR00032. Wales COVID-19 Evidence Centre. A rapid review of the effectiveness of remote consultations versus face-to-face consultations in secondary care surgical outpatient settings. August, 2022.

**This report can be accessed from the WCEC library:**

https://healthandcareresearchwales.org/wales-covid-19-evidence-centre-report-library

**Disclaimer:** The views expressed in this publication are those of the authors, not necessarily Health and Care Research Wales. The WCEC and authors of this work declare that they have no conflict of interest.

**TOPLINE SUMMARY**

### What is a Rapid Review?

Our rapid reviews use a variation of the systematic review approach, abbreviating or omitting some components to generate the evidence to inform stakeholders promptly whilst maintaining attention to bias. They follow the methodological recommendations and minimum standards for conducting and reporting rapid reviews, including a structured protocol, systematic search, screening, data extraction, critical appraisal, and evidence synthesis to answer a specific question and identify key research gaps. They take 1-2 months, depending on the breadth and complexity of the research topic/ question(s), extent of the evidence base, and type of analysis required for synthesis.

### Who is this summary for?

The question was refined following stakeholder consultation from questions about remote consulting suggested by the All Wales Medical Directors, Aneurin Bevan Health Board and the Royal College of Podiatry.

### Background / Aim of Rapid Review

The use of remote consultations and telemedicine approaches significantly increased over the pandemic. There is evidence that some patients still prefer this mode of care delivery and time saving may also enable additional consultations and help to reduce waiting lists. However, the effectiveness of remote consulting for certain specialities, such as surgery, is unclear. We aimed to investigate the effectiveness of video or telephone consultations, particularly focusing on clinical, patient reported and safety outcomes, in adult secondary surgical outpatient care during the COVID-19 pandemic.

### Key Findings

#### Extent of the evidence base

- **3 Prospective cohort studies**
  - 2 conducted in the USA (thyroid/parathyroid and carpal tunnel release surgery)
  - 1 in Chile (abdominal surgery)
- **11 Retrospective cohort studies**
  - 8 conducted in the USA (orthopaedic surgery (n=2); spinal surgery (n=3), cancer-related surgery (n=2), laryngology (n=1))
  - 3 in the UK (orthopaedic surgery (n=2), circumcision (n=1))
- **Patient eligibility varied:** Patient or surgeon preference, decision tool or was not described
- **Numbers of participants varied:** (n=32-535)
- **Consultations varied**: initial assessment, pre and post-op; video or telephone
- **Cohorts studied varied**: one, two or three cohorts
- **Outcomes varied**: conversion to in-person consultation; postoperative complications and attendances; morbidity and mortality; diagnostic agreement; change in management plan; costs

#### Recency of the evidence base

- All studies were published **2021/22**

#### Evidence of effectiveness

Prospective studies

- Post-operative complications were **similar for** telemedicine compared to in-person consultations for **thyroid/ parathyroid and abdominal surgery**.
- For patients undergoing **abdominal surgery, postoperative morbidity** and the need for **additional A&E or in-person visits** were **similar** regardless of the mode of consultation (telemedicine compared to in-person). Additionally, **no postoperative mortality** was reported for either group.
- There was **diagnostic agreement** for **carpal tunnel syndrome patients** from the initial remote consultation and later in-person examination, with **no patients needing a change in management plan**.

Retrospective studies

- **Day-of-surgery cancellation rates** were similar when **pre-anaesthesia evaluations** were conducted via **video-conferencing** compared to in-person for patients scheduled for **cancer-related surgery**.
- Post-operative **readmission and mortality within 30 and 90 days** following **cancer-related surgery** were **similar** regardless of the mode of consultation (telemedicine compared to in-person).
- **Surgical plans** generated via telemedicine for **orthopaedic patients rarely changed** by in-person evaluation.
- There was **mixed** evidence for the **effectiveness of telephone consultations** compared to in-person consultations for orthopaedic patients based on a clinical letter scoring tool.
- **Postoperative complication rates, postoperative visits and reoperation rates** following **orthopaedic surgery** were **similar** regardless of the mode of consultation (telemedicine and telephone calls compared to in-person).
- **Readmission and reoperation rates** were similar when **pre-operative consultations were** conducted via **video-conferencing** compared to in-person for patients scheduled for **spinal surgery;** and video-conferencing also **generated accurate spine surgical plans** that **did not need to change** on the day of surgery.
- **Costs** can be saved, and **time to surgery** is decreased when **pre-operative consultations** are conducted via **telephone calls** compared to in-person for patients undergoing assessment for **circumcision; clinical cancellation rates** were similar for both groups.
- Telemedicine can be used to provide a **preliminary diagnosis and management plan** for **laryngology-related complaints**.

### Policy Implications

- Evidence is of **low quality** but suggests that for many surgical outpatient consultations, **remote consultations are as effective as in-person consultations**.
- There is **potential for time and cost savings** for remote consultations compared to in-person consultations.
- **High quality research is needed** to evaluate the effectiveness of remote consultations to understand which patients and which surgical specialities would benefit most.

### Strength of Evidence

Evidence is **low or very-low quality** due to **observational study designs, small sample sizes and patient selection**.

## Abbreviations

APP: Advanced practice professional
ASA: American Society of Anaesthesiologists Physical Status Classifications
CI: Confidence interval
CTR: Carpal tunnel release
CTS: Carpal tunnel syndrome
CTS-6: Carpal tunnel syndrome 6-item evaluation tool
ED: Emergency department
GRADE: Grading of Recommendations Assessment, Development and Evaluation
IP: In-person
IPC: In-person cohort
NHS: National Health Service
OR: Odd ratio
PAT: Pre-anaesthesia testing
POV: Postoperative visit
PRISMA: Preferred Reporting Items for Systematic Reviews and Meta-Analyses
SD: Standard deviation
SIGN: Scottish Intercollegiate Guidelines Network
TeC: Telephone cohort
TM: Telemedicine
TMC: Telemedicine cohort
UK: United Kingdom
USA: Unites States of America
WHO: World Health Organisation

## 5. RAPID REVIEW METHODS

### 5.1 Eligibility criteria

Inclusion criteria were informed by the PICOS (Population, Intervention, Comparison, Outcome, Study design) framework. Inclusion criteria were also limited to high income countries, as research findings from low- and middle-income countries might not have been fully transferable to the UK context. To check countries income status, World Population Review website (https://worldpopulationreview.com/country-rankings/high-income-countries) was used.

|  | **Include** | **Exclude** |
| --- | --- | --- |
| **Population** | Adults | Children |
| **Intervention** | Video consultations<br>Telephone consultation<br>Initial consultation<br>Follow up consultations (Ongoing management)<br>Pre-operative<br>Postoperative<br>Consultations that affect waiting lists (may include other clinical decision-makers within the multipole disciplinary team e.g. Physios, Nurse Practitioners) | Asynchronous modalities<br>Intraoperative<br>Telerehabilitation<br>Telemonitoring<br>Decentralised services using outreach clinics or remote medical centres<br>Patients video conferencing with a specialist from local healthcare facilities with the aid of local primary care providers<br>Patients having scans (e.g., x-ray) locally and then videoconferencing with an orthopaedic specialist at a later date (virtual fracture clinics) |
| **Comparison** | Face-to-face consultations |  |
| **Outcome** | Waiting list times<br>Safety (Clinical outcomes)<br>- Accuracy of examination<br>- Missed/delayed diagnosis<br>- Change of surgical plan<br>- Postoperative complication rates<br>- Healthcare utilisation (including follow up face-to-face consultations, readmissions, reoperations, emergency department admissions)<br>- Mortality<br>Patient reported outcome measures<br>Costs | Patient satisfaction<br>Patient preferences<br>Provider satisfaction<br>Provider preferences<br>Compliance<br>Barriers and facilitators<br>Surgical / intraoperative outcomes<br>Length of stay<br>Multivariate analysis of usage / people who turn up by a range of demographic / socioeconomic factors |
| **Setting** | All surgical secondary care outpatient settings | Dentistry<br>Maternity |
|  |  | Prison |
| **Study design** | Quantitative<br>- Experimental studies<br>- Prospective observational studies with a comparison group<br>- Prospective and retrospective cohort studies with a comparison group | Qualitative<br>Delphi / consensus studies<br>Cross sectional studies<br>Prospective and retrospective studies without a comparison group |
| **Time frame** | Intervention conducted during or after the COVID pandemic (February 2021 onwards)<br>Dates of search 2021 to 2022 | All cohorts conducted pre COVID |
| **Geographical limitations** | High income countries | Low and middle income countries |

### 5.2 Literature search

Searches were conducted across 6 databases: MEDLINE (on the OVID platform), Embase (on the OVID platform), CINAHL (on the EBSCO platform); WHO Global Coronavirus Database (primary studies), L*OVE COVID (primary studies), Cochrane COVID-19 Study Register, from March 2020 to May 2022 for English language citations.

An initial search of PUBMED was undertaken as part of a rapid evidence summary (May 2022) that informed this rapid review. This was then followed by analysis of the text words contained in the title and abstract, and of the index terms used to describe each article which informed the development of a comprehensive search strategy tailored for each information source. The full search strategies across all the databases are detailed in the additional material. The reference list of all included studies was screened for additional studies. Moreover, forward citation searches for all included studies were conducted with the use of Google Scholar to see whether new publications, that cited the included research papers, could be identified.

All records retrieved from the database searches were imported or entered manually into the refence manager software EndNote X20™ and duplicates removed. Irrelevant records were removed by searching for keywords within the title using the search feature within the Endnote software. The project team agreed which keywords to use to identify papers which did not meet the inclusion criteria. At the end of this process the records that remained were exported as an XML file and then imported to the systematic review software package Rayan™.

### 5.3 Study selection process

The records were screened by a single reviewer with keyword categories for include, exclude highlighted using the Rayyan™. Two reviewers dual screened at least 20% of records using the information provided in the title and abstract resolving all conflicts when needed.

For records that appeared to meet the inclusion criteria, or in cases in which a definite decision could not be made based on the title and/or abstract alone, the full texts of all records were retrieved. Full-text documents were checked by a single reviewer with a screening tool developed for this rapid review containing questions about the inclusion criteria. The screening tool had been piloted on full-text documents found during initial searches, and changes had been made when necessary to make the screening tool fit for purpose. A second reviewer double checked the full-text documents and made a final decision. The flow of records through each stage of the review process will be displayed in a PRISMA flowchart (Page et al. 2021).

### 5.4 Data extraction

All demographic data were extracted directly into tables by one reviewer and checked by another this was piloted on manuscripts for each of the included study designs. The data extracted included specific details about the populations, study methods and outcomes of significance to the review question and specific objectives.

### 5.5 Quality appraisal

The methodological quality of all the research studies were assessed by one reviewer (and judgements verified by a second reviewer) using the JBI critical appraisal checklist for randomised controlled trials (Tufanaru et al. 2020) and the JBI critical appraisal tool for cohort studies (Moola et al. 2020). When a study meets a criterion for inclusion a score of one will be given. Where a particular point for inclusion is regarded as “unclear” it will be given a score of zero. Where a particular point for inclusion is regarded as “not applicable” this point will be taken off the total score. Overall critical appraisal scores will be presented.

Retrospective cohort studies were appraised using the Scottish Intercollegiate Guidelines Network, Methodology Checklist 3; Cohort Studies (Scottish Intercollegiate Guidelines Network 2019). This is a 14-item checklist (‘yes’, ‘no’, ‘can’t say’, ‘does not apply’). Five items do not apply to this type of study design (Statement 1.3, 1,4, 1.5, 1.6, 1.12). Additionally, when there is only one group, statement 1.8 (the assessment of outcome is made blind to exposure status) does not apply and when measures used are completely objective, statement 1.11 (evidence from other sources is used to demonstrate that the method of outcome assessment is valid and reliable) does not apply. The overall assessment reflects how well the study has sought to minimise the risk of bias or confounders. The final rating is high quality (++), acceptable (+) or low quality (-):

- High quality (++): Majority of criteria met. Little or no risk of bias. Results unlikely to be changed by further research
- Acceptable (+): Most criteria met. Some flaws in the study with an associated risk of bias, Conclusions may change in the light of further studies
- Low quality (-): Either most criteria not met, or significant flaws relating to key aspects of study design. Conclusions likely to change in the light of further studies

As retrospective designs are generally regarded as a weaker design, the authors of the checklist suggest that they should not receive a rating higher than “+”.

### 5.6 Synthesis

The data was reported narratively as a series of thematic summaries for each outcome of interest (Thomas et al. 2017)

### 5.7 Assessment of body of evidence

The strength of findings from the thematic summaries of intervention studies were assessed using the Grading of Recommendations, Assessment, Development and Evaluation (GRADE) approach (Guyatt et al. 2008). Due to heterogeneity of the different participant groups, and interventions outcome data was only available for results that arose from single studies and guidance was followed on undertaking the GRADE for data of this type (Ryan & Hill 2016). As the studies retrieved for this rapid review were observational as opposed to interventional the initial quality of the body of evidence overall starts off as low. When rating the evidence based specifically on study design (in particular, retrospective cohort studies) this led to the ratings for all evidence generated using material from these types of study being downgraded from ‘low quality’ to ‘very low quality’.

## 6. EVIDENCE

### 6.1 Study selection flow chart

The PRISMA flow chart (Page et al. 2021) for the review Is displayed in Figure 1 below

**Figure 1:**
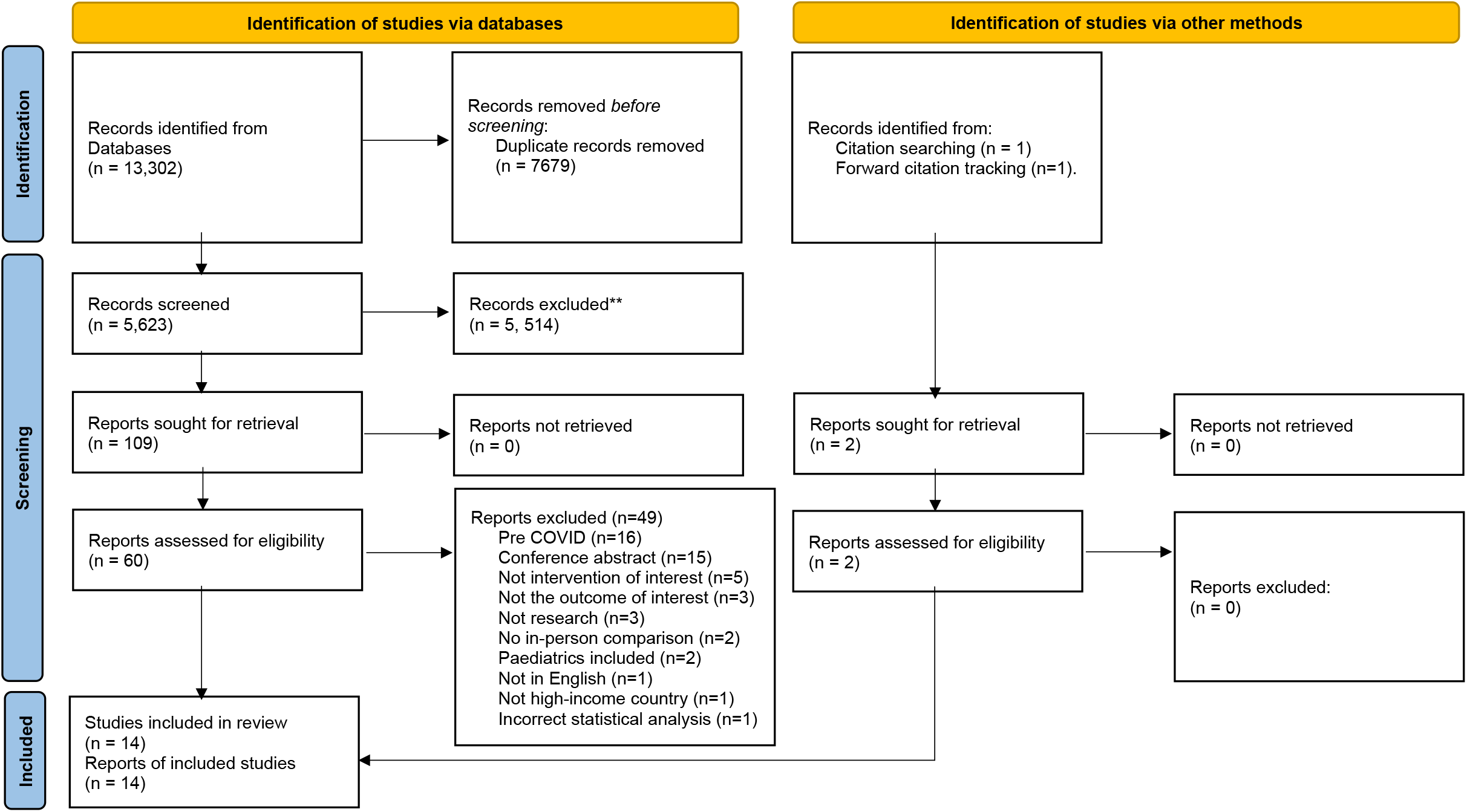
PRISMA flow diagram.

### 6.2 Data extraction tables

The data extraction for the prospective and retrospective studies is displayed in Tables 2 and 3 respectively

**Table 2:** Data extraction for prospective cohort studies.

| Author/s<br>Setting / Country /<br>Aim<br>Details of TM / Patient eligibility | Setting<br>Participant characteristics | Outcomes<br>Data collection methods<br>Critical appraisal score | Relevant findings |
| --- | --- | --- | --- |
| **One cohort** |  |  |  |
| <u>Grandizio et al. 2022</u><br><u>Setting</u><br>Upper extremity surgery division,<br>Level I trauma centre, USA<br><u>Aim</u><br>To compare TM and IP administration of the six item CTS-6 instrument in patients undergoing CTR and to determine whether surgical plans determined via telemedicine were altered by in-person assessments<br>To assess agreement between telemedicine and in-person examinations<br><u>Details of TM</u><br>Initial consultation<br>Video consultation - InTouch Health<br>Conducted by one upper extremity surgeon<br><u>Patient eligibility for TM</u><br>Patient preference<br>TM visits were typically 1 to 3 weeks earlier than the IP visits | <u>Participants</u><br>Patients referred to a hand surgeon for evaluation of CTS (n=32)<br><u>Surgical sub-speciality</u><br>Upper extremity surgery<br><u>Cohorts</u><br>One cohort at two time points seen initially via TM and then IP<br>During COVID<br>(Sep 2020 to March 1, 2021)<br><u>Age (years)</u><br>Mean±SD: 46±12,<br>Range: 24-72<br><u>Gender</u><br>Female (75%)<br><u>Ethnicity</u><br>Not reported<br><u>ASA</u><br>Not reported | <u>Primary outcome/s</u><br>Diagnostic agreement (at initial consultation)<br>Change in management plan<br><u>Outcome measures</u><br>Management plan status (no change, change)<br>Mean CTS-6 score<br>% Agreement<br><u>Data collection methods</u><br>Modified CTS-6 instrument<br>A CTS-6 score > 12 (80% probability of CTS ) was considered diagnostic for CTS<br>- Median nerve numbness<br>- Nocturnal numbness<br>- Thenar atrophy or weakness<br>- Positive Phalen's test<br>- Median nerve sensory changes<br>- Positive Tinel sign<br>Additional tests and manoeuvres<br>- Median nerve compression test (Durkan)<br>- Tinel sign (ulnar nerve at elbow)<br>- Elbow flexion test | <u>Diagnostic agreement</u><br>Mean CTS-6 score<br>TM: 17.7±3.5: IP: 16.8±3.8: p=0.34<br><u>Change in management plan</u><br>There were no cases indicated for CTR during the TM visit that had a subsequent change in management based on the in-person evaluation<br><u>% Agreements</u><br>Median nerve numbness (100%); Kappa 1.00<br>Nocturnal numbness (100%); Kappa 1.00<br>Thenar atrophy or weakness (94%); Kappa 0.88<br>Positive Phalen's test (97%); Kappa 0.94<br>Median nerve sensory changes (63%); Kappa 0.26<br>Positive Tinel sign (78%); Kappa 0.56<br>Median nerve compression test (94%); Kappa 0.88<br>Tinel sign (84%); Kappa 0.68<br>Elbow flexion test (97%); Kappa 0.94 |
|  |  | <u>Critical appraisal score</u><br>6 out of 9<br>JBI critical appraisal checklist for cohort studies (prospective) |  |
| **Two cohorts** |  |  |  |
| <u>Boles et al 2022</u><br><u>Setting</u><br>Tertiary care centre, USA<br><u>Aim</u><br>To compare the safety and efficacy of telemedicine with in-person pre-operative visits in patients undergoing thyroid and parathyroid surgery<br><u>Details of TM</u><br>Pre- and postoperative consultations<br>No further details reported<br><u>Patient eligibility for TM</u><br>All new patients were offered an initial telemedicine consultation but were given options for in-person appointment by request<br>Surgeon directed conversion to IP evaluation was based on clinical judgement and was especially considered for patients with previous central neck surgery, history of voice or swallowing changes, or signs of high-risk thyroid cancer such as lymph node metastases or extra-thyroidal extension | <u>Participants</u><br>Patients undergoing thyroid and parathyroid surgery (n=94)<br><u>Cohorts</u><br>Two cohorts<br>TMC: n=28<br>IPC: n=66<br>During COVID<br>(March to Oct 2020)<br><u>Age (years)</u> Mean±SD<br>TMC: 47.1±16.4; IPC: 53.3±18.1<br><u>Gender</u><br>Female: TMC: 78.6%; IPC: 68.2%<br><u>Ethnicity</u><br>White: TMC:39.3%; IPC: 39.4%<br>Hispanic: TMC: 35.7%; IPC: 22.7%<br>Black: TMC: 0%; IPC: 1.5%<br>Asian: TMC: 14.3%; IPC: 10.6%<br>Other: TMC: 10.7%; IPC: 25.8%<br><u>ASA</u><br>Not reported | <u>Primary outcome/s</u><br>Conversion from TM to IP consultations<br>Postoperative complications<br>- Persistent hypocalcaemia<br>- Recurrent laryngeal nerve paralysis<br>- Laryngeal paresis<br>- Dysphonia<br>- Transient voice changes<br>- Swallow changes<br>- Postoperative infections<br>- General medical complications<br>- Readmissions<br><u>Outcome measures</u><br>Conversion to in-person consultation rates<br>Postoperative complication rates<br><u>Data collection methods</u><br>REDCap electronic data capture tools<br><u>Critical appraisal score</u><br>7 out of 11<br>JBI critical appraisal checklist for cohort studies (prospective) | <u>Conversion to in-person consultation</u><br>Six patients had their pre-operative visit converted from telemedicine to in-person for a variety of reasons<br>Regarding postoperative follow up, most patients opted for telemedicine visits, with 61 patients seen via telemedicine, 30 patients seen in-person, and three had their postoperative visit converted from telemedicine to in-person<br>More than half (n=37, 56%) of the IPC opted to utilize TM for postoperative follow up<br><u>Postoperative complication rates</u><br>TMC: 7.1% vs IPC: 9.1%; p=1.00 |
| <u>Irarrázaval et al. 2021</u><br><u>Setting</u> | <u>Participants</u> | <u>Primary outcome/s</u><br>Postoperative morbidity<br>Postoperative mortality | <u>Postoperative morbidity rate</u><br>TMC: 5.7% vs IPC 8%; p=0.50<br><u>Postoperative mortality rate</u> |
| Department of Gastrointestinal Surgery, Chile<br><u>Aim</u><br>To compare the use of TM clinics to IP follow-up for postoperative care after gastrointestinal surgery during COVID-19 outbreak<br><u>Details of TM</u><br>Postoperative consultations<br>Video consultation - bespoke platform<br>Telephone consultation<br>Conducted by an attending surgeon<br><u>Patient eligibility for TM</u><br>Patient preference | All abdominal surgery patients operated since the COVID-19 pandemic (n=219)<br><u>Surgical sub-speciality</u><br>Gastroesophageal, hepatobiliary, colorectal and general surgery procedures<br><u>Cohorts</u><br>Two cohorts<br>TMC & TeC: n=106<br>IPC: n=113<br>During COVID<br>(March 15 to July 19, 2020)<br><u>Age (years) Mean±SD</u><br>TMC: 49±20; IPC: 53±16<br><u>Gender</u><br>Female: TMC: 55%; IPC: 52%<br><u>Ethnicity</u><br>Not reported<br><u>ASA</u><br>ASA 1-2: TMC: 94%; IPC: 96%<br>ASA 3 or more: TMC: 6%; IPC: 4% | Minor complications<br>Major complications<br>Additional in-person visits<br>ED visits<br><u>Outcome measures</u><br>Postoperative morbidity rates<br>Postoperative mortality rates<br>Minor complication rates<br>Major complication rates<br>Numbers of additional in-person visits and ED visits within 30 days of surgery<br><u>Data collection methods</u><br>Prospective database<br><u>Critical appraisal score</u><br>7 out of 11<br>JBI critical appraisal checklist for cohort studies (prospective) | No mortality was reported<br><u>Minor complications rates</u><br>TMC: 6% vs IPC: 8%; p=0.79<br><u>Major complications rates</u><br>TMC: 0% vs IPC: 0.9%; p>0.99<br><u>Additional in-person visits</u><br>2.8% (3/106) of patients in the TMC & TeC required a subsequent in-person<br>3.5% (4/113) of patients in the IPC had a subsequent in-person visit<br>TMC: 2.8% vs IPC: 3.5%; p=0.09<br><u>ED visits</u><br>1.9% (2/106) of patients in the TMC & TeC visited the ED within 30 days after surgery<br>6.2% (7/113) of patients in the IPC visited the ED within 20 days after surgery<br>TMC: 1.9% vs IPC: 6.2%; p=not reported |

Key: ASA: American Society of Anaesthesiologists Physical Status Classifications; ED: emergency department; CTR: carpal tunnel release; CTS: carpal tunnel syndrome; IP: in-person; IPC: in-person cohort; SD: standard deviation; TeC: telephone cohort; TM: telemedicine; TMC: telemedicine cohort

**Table 3:** Data extraction for retrospective cohort studies.

| Author/s<br>Setting / Country /<br>Aim<br>Details of TM / Patient eligibility | Participant characteristics | Outcomes<br>Data collection methods<br>Critical appraisal score | Relevant findings |
| --- | --- | --- | --- |
| **One cohort** |  |  |  |
| <u>Choi et al. 2022</u><br><u>Setting / Country</u><br>Tertiary care centre laryngology clinic, USA<br><u>Aim</u><br>To investigate the concordance in diagnosis and management between initial telemedicine visits and subsequent in-person visits with laryngoscopy for laryngology-related complaints during COVID-19.<br><u>Details of TM</u><br>Initial consultation<br>Video consultation - USC telecare application<br>Conducted with a laryngologist<br><u>Patient eligibility for TM</u><br>Patient preference | <u>Participants</u><br>All new patient referrals with laryngology-related complaints for an initial TM appointment and a subsequent follow up IP with laryngoscopy (n=250)<br><u>Laryngology-related complaints</u><br>Voice (n=128); swallowing (n=23); airway (n=20); general throat complaints (n=54); other (n=25)<br><u>Cohorts</u><br>One cohort<br>During COVID<br>(March 17 to Oct 26, 2020)<br><u>Age (years)</u><br>Mean±SD: 50.1±17<br><u>Gender</u><br>Female: 54.8%<br><u>Ethnicity</u><br>White: 36.4%; Asian: 14.8%<br>Hispanic: 13.2%; Black: 3.2%<br>Other: 32.4%<br><u>ASA</u><br>Not reported | <u>Primary outcome/s</u><br>Concordance between TM and subsequent IP visit with laryngoscopy<br><u>Outcome measures</u><br>Concordance rates diagnosis and management<br><u>Data collection methods</u><br>Retrospective chart review<br><u>Critical appraisal score</u><br>Acceptable based on SIGN methodology checklist 3 for cohort studies (retrospective) | <u>Concordance rates diagnosis</u><br>86.1% (215/250)<br>There were no statistical differences in concordance rates of diagnoses by chief complaint<br>Pre- and post-laryngoscopy diagnoses were rated to be discordant among 35 patients (14%). Of these 19 patients were rated to have concordant management without any additional or different management plans<br><u>Concordance rates management</u><br>93.7% (234/250)<br>The concordance rates in management were significantly lower among patients with general throat complaints (OR : 0.27, 95% CI: 0.08–0.90) in comparison with voice-related complaints<br>When adjusted for patient demographics, provider, and relevant clinic factors, the differences were no longer significant (OR 0.28, 95% CI: 0.06–1.26)<br>16 patients had discordant pre- and post-laryngoscopy diagnosis and management (after laryngoscopy, patients required additional imaging, different procedures, referred to<br>gastroenterologists or was admitted to inpatient from clinic for airway monitoring and tracheostomy) |
| <u>Crawford et al. 2021</u><br><u>Setting</u><br>Departments of orthopaedic surgery at two institutions, USA<br><u>Aim</u><br>To assess whether surgical plans proposed following telemedicine visits changed after subsequent in-person interaction and to explore these changes across subspecialties<br><u>Details of TM</u><br>Pre-operative consultation<br>No details provided<br><u>Patient eligibility for TM</u><br>Not reported | <u>Participants</u><br>Orthopaedic surgical patients indicted for surgery during a virtual visit who had clear and specific surgical plan documented in the medical records and a subsequent in-person, prior to surgery (n=303)<br><u>Orthopaedic sub-speciality</u><br>Arthroplasty (n=77); Foot and ankle (n=11); Spine (n=59); Sports (n=80); Upper extremity (n=76)<br><u>Cohorts</u><br>One cohort<br>During COVID<br>(March 1 to July 31, 2020)<br><u>Age (years)</u><br>Mean±SD: 54±17.5<br><u>Gender</u><br>Female: 45.2%<br><u>Ethnicity</u><br>Not reported<br><u>ASA</u><br>ASA 1: 17.2%<br>ASA 2: 54.8%<br>ASA 3: 27.4%<br>Not reported: 0.7% | <u>Primary outcome/s</u><br>Change in surgical plan<br>- A patient indicated for surgery via telemedicine was not found to warrant surgery after in-person evaluation<br>- The procedure described during the telemedicine encounter was changed, or additional procedures added, following in-person evaluation<br>- Additional procedures were added after in-person evaluation<br><u>Outcome measures</u><br>Surgical plan status<br>(no change, change)<br><u>Data collection methods</u><br>Retrospective chart review<br><u>Critical appraisal score</u><br>Acceptable on SIGN methodology checklist 3 for cohort studies (retrospective) | <u>Change in surgical plan</u><br>In 96% (292/ 303) of cases the surgical plan did not significantly change after in-person evaluation (proportion of change: 0.04, 95% CI 0.02–0.06)<br>By subspecialty, plans remained the same for 77/77 patients (100%) in hip and knee arthroplasty<br>77/80 patients (96%) in sports surgery (proportion of change: 0.04, 95% CI 0.01–0.11)<br>73/76 patients (96%) in upper extremity/shoulder surgery (proportion of change: 0.04, 95% CI 0.01–0.11)<br>54/59 patients (92%) in spine surgery (proportion of change: 0.08, 95% CI 0.03–0.19)<br>11/11 patients (100%) in foot and ankle surgery |
| <u>Lightsey et al. 2021</u><br><u>Setting</u> | <u>Participants</u><br>All new patients who were indicated for an elective spine surgery at the time of a | <u>Primary outcome/s</u><br>Change in surgical plan | <u>Change in surgical plan</u> |
| Division of spine surgery<br>Department of orthopaedic surgery, USA<br><u>Aim</u><br>To characterize the impact of telemedicine on spine surgical planning by assessing whether surgical plans established in virtual visits changed following in-person evaluation<br><u>Details of TM</u><br>Pre-operative consultation<br>Conducted by four surgeons<br>No further details provided<br><u>Patient eligibility for TM</u><br>Exclusions<br>- Patients requiring urgent or emergent assessment and surgery<br>- Those patients with complex presentations requiring in-person evaluation | virtual encounter and subsequently underwent an in-person evaluation prior to the procedure whether in the form of a pre-operative clinic appointment or assessment on day of surgery (n=33)<br><u>Orthopaedic sub-speciality</u><br>Spine surgery (n=33)<br><u>Cohorts</u><br>One cohort<br>During COVID<br>(March 1 to July 31, 2020)<br><u>Age (years)</u><br>Mean±SD: 59.3+14.2<br><u>Gender</u><br>Female: 33.3%<br><u>Ethnicity</u><br>Not reported<br><u>ASA</u><br>ASA 1: 0%<br>ASA 2: 44.6%<br>ASA 3: 50%<br>ASA 4: 3.1% | - Patient previously indicated for surgery was found not to merit a surgical procedure<br>- Change in the type of surgery indicated<br>- Change in the extent of surgery offered<br><u>Outcome measures</u><br>Surgical plan status<br>(no change, change)<br><u>Data collection methods</u><br>Retrospective chart review<br><u>Critical appraisal score</u><br>Acceptable on SIGN methodology checklist 3 for cohort studies (retrospective) | In 94% (31/34) of cases, the pre-operative plan did not significantly change after in-person evaluation<br>(95% CI 1% - 20%) |
| **Two cohorts - during COVID comparison group** |  |  |  |
| <u>Aldawoodi et al. 2021</u><br><u>Setting</u><br>Cancer centre, USA<br><u>Aim</u><br>To demonstrate telemedicine pre-anaesthesia evaluation as a reasonable and convenient option | <u>Participants</u><br>All oncologic surgical patients undergoing pre-anaesthesia evaluation (across a range of surgical specialities) (n=238)<br><u>Surgical sub-speciality</u><br>Breast/plastics (n=52); cutaneous (n=28); head and neck (n=9); gastrointestinal (n=17); urology (n=43); gynaecology (n=60); interventional radiology (n=3); | <u>Primary outcome/s</u><br>Day of surgery cancellations<br><u>Outcome measures</u><br>Cancellation rate<br><u>Data collection methods</u><br>Retrospective chart review<br><u>Critical appraisal score</u> | <u>Cancellation rate</u><br>TMC: 1.67% vs IPC: 0%; p=0.4979 |
| for eligible presurgical oncology patients<br><u>Details of TM</u><br>Pre-anaesthesia evaluation<br>Video consultation - Zoom<br>Conducted with an APP<br><u>Patient eligibility for TM</u><br>PAT decision tool | neurosurgery (n=3); orthopaedic (n=5); pulmonary (n=9); sarcoma (n=8); thoracic (n=1)<br><u>Cohorts</u><br>Two cohorts<br>TMC: n=120<br>During COVID<br>(June 29 to Sept 22, 2020)<br>IPC: n=118/120<br>During COVID<br>(June 3 to June 26, 2020)<br>Patients who would have been eligible for telemedicine evaluation based upon the PAT decision tool, which was applied post hoc based upon chart review<br><u>Age (years)</u><br>Mean<br>TMC: 57; IPC: 68<br><u>Gender</u><br>Female (TMC: 69.2%; IPC: 61.0%)<br><u>Ethnicity</u><br>White (TMC: 85.8%; IPC: 81.4%)<br>Black (TMC: 6.7%; IPC: 9.3%)<br>Asian (TMC: 0.8%; IPC: 0.8%)<br>Pacific Islander (TMC: 0.8%; IPC: 0.8%)<br>Native American (TMC: 0.8%; IPC: 0%)<br>Other (TMC: 4.2%; IPC: 7.6%)<br>Unknown (TMC: 0.8%; IPC: 0%)<br><u>ASA</u><br>ASA 1 (TMC: 0.83%; IPC: 0%)<br>ASA 2 (TMC: 62.5%; IPC: 39%)<br>ASA 3 (TMC: 36.7%; IPC: 60.2%) | Low quality on SIGN methodology checklist 3 for cohort studies (retrospective) |  |
| <u>Natale et al. 2022</u> | <u>Participants</u> | <u>Primary outcome/s</u> | <u>Cancellation Rate</u> |
| <u>Setting</u><br>Department of general surgery, UK<br><u>Aim</u><br>To determine whether standalone teleconsultation is an effective alternative to face-to-face assessment of patients requiring circumcision<br><u>Details of TM</u><br>Pre-operative consultation<br>Telephone calls<br>No further details provided<br><u>Patient eligibility for TM</u><br>No details provided | Patients listed for circumcision (combined procedures or circumcision for penile cancer excluded) (n=101)<br><u>Cohorts</u><br>Two cohorts<br>TeC: n=42<br>IPC: n=59<br>During COVID<br>1 Feb to 30 Sept 2020<br><u>Age (years)</u><br>Mean<br>TeC: 36; IPC: 50<br><u>Gender</u><br>Males (100%)<br><u>Ethnicity</u><br>Not reported<br><u>ASA</u><br>Not reported | Clinical cancellations<br>- Any cancellation related to patient health, operative or anaesthetic factors<br>Time to theatre<br>Costs<br><u>Outcome measures</u><br>Cancellation rate<br>Cost savings<br>Mean time to theatre<br><u>Data collection methods</u><br>Retrospective chart review<br><u>Critical appraisal score</u><br>Acceptable on SIGN methodology checklist 3 for cohort studies (retrospective) | Entire cohort: OR 0.063 (95% CI: 0.024–0.13)<br>The odds of cancellation did not significantly differ between the telephone and face-to-face groups, OR 0.37 (95% CI: 0.039–3.46)<br><u>Time to theatre</u><br>IPC: 181 days (CI: 152-210)<br>TeC: 70 days (CI: 57-82)<br>p<.0.01<br><u>Cost Savings</u><br>Per-appointment cost:<br>IPC: £241.36<br>TeC: £102.56<br>The overall cost of clinic assessment for the cohort was £18,547.76. If all patients were assessed in a telephone clinic, a cost saving of £8189.20 could have been achieved |
| <u>Uppal et al. 2022</u><br><u>Setting</u><br>Cancer centre, USA<br><u>Aim</u><br>To measure short-term outcomes of patients with postoperative telemedicine visits compared with in-person visits<br><u>Details of TM</u><br>Postoperative consultation<br>Telemedicine POV was defined as the use of a virtual, video-, or | <u>Participants</u><br>All patients undergoing elective inpatient cancer-related surgery for a POV visit (n=535)<br><u>Primary surgical service</u><br>Colorectal (n=245), pancreas (n=39), liver (n=133), gastric, peritoneal cytoreduction, and sarcoma (n=159) surgical procedures<br><u>Cohorts</u><br>Two cohorts<br>TMC: n=98<br>IPC: n=437<br>During COVID<br>(March to Dec 2020) | <u>Primary outcome/s</u><br>90-day readmissions<br><u>Secondary outcome/s of interest</u><br>30-day readmissions<br>Mortality<br><u>Outcome measures</u><br>90 day readmission rates<br>30 day readmission rates<br>90 day mortality rates<br>Median time to readmission<br><u>Data collection methods</u><br>Retrospective chart review | <u>Readmission rates</u><br>A total of 60 (11.2%) of patients were readmitted within 90 days of discharge from their operative hospitalization.<br><u>Median time to readmission</u><br>TMC: 21 days vs IPC: 17 days; p=0 .585<br><u>30-day readmission rate</u><br>TMC: 7.1% vs IPC: 11.4%; p=0.29<br><u>90-day readmission rate</u><br>TMC: 16.3% vs IPC: 16.5%; p=0.99<br><u>90-day mortality rate</u><br>TMC: 0%; IPC: 0.20%; p=1.00 |
| audio-based encounter (if the patient was unable to use their video camera) for the first appointment following discharge after surgery<br>No further details provided<br><u>Patient eligibility for TM</u><br>The decision for in-person or telemedicine POV was at the discretion of the primary surgeon and the patient, and determined before discharge | <u>Age (years) Mean±SD</u><br>TMC: 56.6±14.0; IPC 59.1±12.81<br><u>Gender</u><br>Female: TMC: 46.9%; IPC: 42.6%<br><u>Ethnicity</u><br>Asian: TMC: 3.31%; IPC: 4.1%<br>Black or African American: TMC: 7.1%; IPC: 9.2%<br>Other: TMC: 3.1%; IPC: 9.8%<br>White or Caucasian: TMC: 86.7%; IPC: 76.9%<br><u>ASA</u><br>Not reported | <u>Critical appraisal score</u><br>Acceptable on SIGN methodology checklist 3 for cohort studies (retrospective) | Reasons for readmission within 90 days did not differ between patients in either cohort |
| **Two cohorts - pre COVID comparison group** |  |  |  |
| <u>Greven et al. 2022</u><br><u>Setting</u><br>Spine centre, USA<br><u>Aim</u><br>To evaluate the safety and efficacy of using telemedicine alone to preoperatively evaluate spine surgery<br><u>Details of TM</u><br>Pre-operative consultation<br>Conducted by the neurosurgical and anaesthesia teams<br>No further details provided<br><u>Patient eligibility for TM</u><br>If there was any question of appropriateness of the procedure, safety, or high-risk status of a | <u>Participants</u><br>All spine surgery patients seen by 3 neurological spine surgeons (n=276)<br><u>Spine segment involved</u><br>Cervical: n=88; Thoracic: n=16<br>Lumbar: n=160; Sacral: n=2<br><u>Cohorts</u><br>Two cohorts<br>TMC: n=138<br>During COVID<br>(April 1 to Sep 15, 2020)<br>IPC: n=138<br>Pre COVID<br>(April 1 to Sep 15, 2019)<br><u>Age (years) Mean±SD</u><br>TMC: 60.2±14.3; IPC: 61.4±13.7<br><u>Gender</u> | <u>Outcome/s of interest</u><br>Readmissions<br>Reoperations<br><u>Outcome measures</u><br>Readmission rate<br>Reoperation rate<br><u>Data collection methods</u><br>Retrospective chart review<br><u>Critical appraisal score</u><br>Low quality on SIGN methodology checklist 3 for cohort studies (retrospective) | <u>Readmission rate</u><br>TMC: 7.9%; IPC 4.3%: p=0.208<br><u>Reoperation rate</u><br>TMC: 10.1%; IPC 5.1%: p=0.091 |
| patient, an in-person visit was scheduled prior to surgery | Female: TMC: 45%; IPC: 53%<br><u>Ethnicity</u><br>Not reported<br><u>ASA</u><br>Mean score<br>TMC: 2.6±0.6; IPC: 61.4±13.7 |  |  |
| <u>Henry et al. 2022</u><br><u>Setting</u><br>Orthopaedic practice, USA<br><u>Aim</u><br>To assess postoperative healthcare utilization in patients seen via telehealth for at least one clinical visit after upper extremity surgery and determine the conversion rate to in-person evaluation<br><u>Details of TM</u><br>Postoperative consultation<br>Video consultation (91.7%) - platform not specified<br>Telephone consultation (8.3%)<br>Conducted by one of 12 fellowship-trained upper extremity surgeons<br><u>Patient eligibility for TM</u><br>Treating surgeons chose which patients would be seen via telemedicine | <u>Participants</u><br>All orthopaedic patients seen for a postoperative telemedicine visit (n=112)<br><u>Orthopaedic sub-speciality</u><br>Hand and upper extremity surgery<br><u>Cohorts</u><br>Two cohorts<br>TMC & TeC: n=56 (60 visits)<br>During COVID<br>(April and May 2020)<br>IPC: n=56<br>Pre COVID<br>(Matched surgical procedures between 2018 and 2020)<br><u>Age (years) Mean±SD</u><br>TMC & TeC: 59±14.8; IPC: 60<br><u>Gender</u><br>Female: TMC & TeC: 57%; IPC: 66%<br><u>Ethnicity</u><br>Not reported<br><u>ASA</u><br>Not reported | <u>Primary outcome/s</u><br>Conversion to in-person evaluation<br>Changes to the postoperative treatment course<br>Number of postoperative visits<br><u>Secondary outcome/s</u><br>Postoperative complications<br>Reoperations<br><u>Outcome measures</u><br>- Conversions to in-person evaluation rate<br>- Change to treatment plan status<br>(Change, no change)<br>- Mean number of postoperative visits<br>- Postoperative complication rate<br>- Reoperation rate<br><u>Data collection methods</u><br>Retrospective chart review<br><u>Critical appraisal score</u><br>Acceptable on SIGN methodology checklist 3 for cohort studies (retrospective) | <u>Conversion to in-person evaluation</u><br>One (1.7%) telemedicine visit required conversion to in-person evaluation due to suspected superficial infection necessitating an in-depth physical examination<br><u>Changes to the postoperative treatment course</u><br>A specific change to the postoperative treatment course solely based on the findings of the telemedicine visit was made in 5.0% (3/112) of cases<br><u>Mean number of postoperative visits</u><br>TMC & TeC (2.6 visits; range: 1-7 visits) versus IPC (2.7 visits; range: 1-6 visits); p=0.886<br><u>Postoperative complication rate</u><br>TMC & TeC 3.6% versus IPC 7.1%; p=0.679<br><u>Reoperation rate</u><br>TMC & TeC 0.0% versus IPC 1.8%; p=1.00 |
| <u>Raad et al 2021</u> | <u>Participants</u> | <u>Primary outcome/s</u> | <u>Effectiveness of consultations</u> |
| <u>Setting</u><br>Trauma and orthopaedic clinic, UK<br><u>Aim</u><br>To determine the efficacy of telephone medicine consultations in trauma and orthopaedics<br><u>Details of TM</u><br>Initial and postoperative consultations<br>Telephone calls<br>No further details provided<br><u>Patient eligibility for TM</u><br>No further details provided | All new referrals and follow-up orthopaedic (operative and non-operative) patients (n=180)<br><u>Orthopaedic sub-speciality</u><br>Not reported<br><u>Cohorts</u><br>Three cohorts (two TeC cohorts were combined for analysis)<br>TeC1: n=60<br>During COVID: April 2020<br>TeC2: n=60<br>During COVID: May 2020<br>IPC: n=60<br>Pre COVID: March 2020<br>No further participant details provided | Effectiveness of consultations (quality and efficacy of consultations)<br>- Diagnosis<br>- Investigations<br>- Treatment plan<br>- Value of the consultation<br><u>Outcome measures</u><br>Ashford Clinic Letter Scoring system<br>Total score out of 8<br><u>Data collection methods</u><br>Retrospective chart review (clinic letters)<br><u>Critical appraisal score</u><br>Low quality on SIGN methodology checklist 3 for cohort studies (retrospective) | Mean overall score (new patients and follow-up) IPC: 6.7±1.15; TeC1 & 2: 7.275±0.66 (p<0.001)<br>The relative risk of failing to make a diagnosis with a telephone consultation as compared to a physical appointment was 0.388 (95% CI 0.14-1.07) |
| <u>Ye et al. 2022</u><br><u>Setting</u><br>Department of orthopaedic surgery, USA<br><u>Aim</u><br>To compare accuracy of surgical plans generated from in-person and telemedicine evaluations and assess the reasons for surgical plan changes between initial evaluation and surgery<br>The secondary objective was to assess the effect of changes in | <u>Participants</u><br>New patients scheduled for orthopaedic spine surgery where a surgeon documented a definitive surgical plan at the initial visit. (n=131)<br><u>Spine procedures performed</u><br>Anterior cervical (n=28), posterior cervical (n=15), lumbar decompression (n=50), posterior lumbar fusion (n=38)<br><u>Cohorts</u><br>Two cohorts<br>TMC: n=39<br>During COVID (April 2020 to Oct 2020) | <u>Primary outcome/s of interest</u><br>Change in surgical plan<br>- A change in the extent of surgery offered<br>- A change in the approach<br>- A change in type of surgery<br>- A change in region of surgery<br>Time to surgery<br><u>Outcome measures</u><br>Surgical plan status (no change, change)<br>Mean days to surgery<br><u>Data collection methods</u><br>Retrospective chart review | <u>Change in surgical plan</u><br>TMC: 79.5% vs IPC: 82.6%; p=0.673<br>There was no difference in hospital complication rate (p=0.461), 30-day readmission (p= 0.726), or 6-month reoperation p=0.921) between patients with consistent and non-consistent surgical plans<br><u>Days to surgery</u><br>The telemedicine cohort experienced significantly longer time between the initial appointment and surgery (44 days vs. 33 days, p=0.002) compared with the in-person cohort |
| surgical planning on postoperative outcomes<br><u>Details of TM</u><br>Pre-operative consultation<br>No further details provided<br><u>Patient eligibility for TM</u><br>No details reported | IPC: n=92<br>Pre COVID<br>(Jan 2019 to July 2019)<br><u>Age (years)</u> Mean±SD<br>TMC: 52.7±13.5<br>IPC: 55.8±13.8<br><u>Gender</u><br>Female: TMC: 59.0%; IPC: 43.5%<br><u>Ethnicity</u><br>White: TMC: 64.1%; IPC: 76.1%<br>Black: TMC: 28.2%; IPC: 22.8%<br>Other: 7.7%; IPC: 1.1%<br><u>ASA</u><br>ASA 1 (TMC: 7.89%; IPC: 1.16%)<br>ASA 2 (TMC: 47.4%; IPC: 43.0%)<br>ASA 3 (TMC: 42.1%; IPC: 54.7%)<br>ASA 4 (TMC: 2.63%; IPC: 1.16%) | <u>Critical appraisal score</u><br>Low quality on SIGN methodology checklist 3 for cohort studies (retrospective) |  |
| **Three cohorts – during COVID comparison groups** |  |  |  |
| <u>Sibanda et al. 2021</u><br><u>Setting</u><br>Shoulder and elbow outpatient clinics, UK<br><u>Aim</u><br>To assess and compare the effectiveness of consultations, that is, telephone, video, and face-to-face in a shoulder and elbow clinic<br><u>Details of TM</u><br>Initial and postoperative consultations | <u>Participants</u><br>New referrals and follow up orthopaedic patients (n=84)<br><u>Orthopaedic sub-speciality</u><br>Shoulder and elbow<br><u>Cohorts</u><br>Three cohorts<br>IPC: n=30<br>TeC: n=30<br>TMC: n=24<br>During COVID<br>(March to April 2021)<br>No further participant details provided | <u>Primary outcome/s</u><br>Effectiveness of consultations (quality and efficacy of consultations)<br>- Diagnosis<br>- Investigations<br>- Treatment plan<br>- Value of the consultation<br><u>Outcome measures</u><br>Ashford Clinic Letter Scoring system<br>Total score out of 8<br><u>Data collection methods</u><br>Retrospective chart review (clinic letters) | <u>Effectiveness of consultations</u><br>Mean overall score (new patients and follow-up)<br>IPC: 7.967, TMC: 7.667; TeC: 7.333 (p=0.0091)<br>IPC versus TeC (p<0.05)<br>IPC versus TMC (p=0.33)<br>TeC versus TMC (p=0.25) |
| Videoconference: accuRx and Attend Anywhere software<br>Telephone calls<br>Conducted with consultants, middle-grade registrars and clinical fellows<br><u>Patient eligibility for TM</u><br>No further details provided |  | <u>Critical appraisal score</u><br>Low quality on SIGN methodology checklist 3 for cohort studies (retrospective) |  |

Key: APP: advanced practice professional; ASA: American Society of Anaesthesiologists Physical Status Classifications; CI: confidence interval; IP: in-person; IPC: in-person cohort; OR: odds ratio; PAT: pre-anaesthesia testing; POV: postoperative visit; SD: standard deviation; SIGN: Scottish Intercollegiate Guidelines Network; TeC: telephone cohort; TM: telemedicine; TMC: telemedicine cohort

### 6.3 Additional material

1. Full search strategies
2. Critical appraisal scores
3. Excluded studies
4. GRADE evidence profile

This is available at: http://www.primecentre.wales/resources/Supplementary/Wales_COVID-19_Evidence_Centre_remote_consultations_versus_face-to-face_consultations_in_secondary_care_surgical_outpatient_settings_Additional_material_july_2022.pdf

## 7.2 Acknowledgements

The authors would like to thank David Charles Bosanquet, Professor Cathy Holt, Sally Rees, Jennifer Wong-Cheetham and Deb Smith for their contribution in guiding the focus of the review.

Key: ASA: American Society of Anaesthesiologists Physical Status Classifications; ED: emergency department; CTR: carpal tunnel release; CTS: carpal tunnel syndrome; IP: in-person; IPC: in-person cohort; SD: standard deviation; TeC: telephone cohort; TM: telemedicine; TMC: telemedicine cohort

## 8. ABOUT THE WALES COVID-19 EVIDENCE CENTRE (WCEC)

The WCEC integrates with worldwide efforts to synthesise and mobilise knowledge from research.

We operate with a core team as part of Health and Care Research Wales, are hosted in the [Wales Centre for Primary and Emergency Care Research (PRIME)](http://www.primecentre.wales/), and are led by [Professor Adrian Edwards of Cardiff University](https://www.cardiff.ac.uk/people/view/123022-edwards-adrian).

The core team of the centre works closely with collaborating partners in [Health Technology Wales](https://www.healthtechnology.wales/), Wales Centre for Evidence-Based Care, [Specialist Unit for Review Evidence centre](https://www.cardiff.ac.uk/specialist-unit-for-review-evidence), [SAIL Databank](https://healthandcareresearchwales.org/about-research-community/secure-anonymised-information-linkage-sail-databank), Bangor Institute for Health & Medical Research/ Health and Care Economics Cymru, and the [Public Health Wales Observatory](https://phw.nhs.wales/services-and-teams/observatory/).

Together we aim to provide around 50 reviews per year, answering the priority questions for policy and practice in Wales as we meet the demands of the pandemic and its impacts.

### Director

Professor Adrian Edwards

### Website

https://healthandcareresearchwales.org/about-research-community/wales-covid-19-evidence-centre

## REFERENCES

Aldawoodi NN, Muncey AR, Serdiuk AA, et al. (2021). A retrospective analysis of patients undergoing telemedicine evaluation in the preanesthesia testing clinic at H. Lee Moffitt cancer center. Cancer control. 28: 10732748211044347. doi: https://doi.org/10.1177/10732748211044347

Boles RW, Zheng M, Kwon D. (2022). Expanded use of telemedicine for thyroid and parathyroid surgery in the COVID-19 era and beyond. American Journal of Otolaryngology. 43(2): 103393. doi: https://doi.org/10.1016%2Fj.amjoto.2022.103393

Cabrera CI, Ning AY, Cai Y, et al. (2020). Systematic review of telehealth cost minimization for patients and health systems in otolaryngology. The Laryngoscope. 131(8): 1741–8. doi: https://doi.org/10.1002/lary.29321

Chaudhry H, Nadeem S, Mundi R. (2021). How satisfied are patients and surgeons with telemedicine in orthopaedic care during the COVID-19 pandemic? a systematic review and meta-analysis. Clinical Orthopaedics and Related Research. 479(1): 47–56. doi: 10.1097/corr.0000000000001494

Choi JS, Yin V, Wu F, et al. (2022). Utility of telemedicine for diagnosis and management of laryngology-related complaints during COVID-19. The Laryngoscope. 132(4): 831–7. doi: https://doi.org/10.1002/lary.29838

Crawford AM, Lightsey HM, Xiong GX, et al. (2021). Telemedicine visits generate accurate surgical plans across orthopaedic subspecialties. Archives of Orthopaedic and Trauma Surgery. Apr 18;1–8.(Online ahead of print).

Fahey E, Elsheikh MFH, Davey MS, et al. (2021). Telemedicine in orthopedic surgery: a systematic review of current evidence. Telemedicine and e-Health. Aug 10 (onlline ahead of print). doi: https://doi.org/10.1089/tmj.2021.0221

Gachabayov M, Latifi LA, Parsikia A, et al. (2022). The role of telemedicine in surgical specialties during the COVID-19 pandemic: a scoping review. World Journal of Surgery. 46(1): 10–8. doi: https://doi.org/10.1007/s00268-021-06348-1

Grandizio LC, Barreto Rocha DF, Foster BK. (2022). Evaluation of a comprehensive telemedicine pathway for carpal tunnel syndrome: a comparison of virtual and in-person assessments. The Journal of Hand Surgery. 47(2): 111–9. doi: https://doi.org/10.1016/j.jhsa.2021.08.024

Greven ACM, McGinley BM, Nakirikanti AS, et al. (2022). Telemedicine in spine surgery: outcomes for 138 patients with virtual preoperative assessment compared to historical controls. World neurosurgery. 161: e495–e9. doi: https://doi.org/10.1016/j.wneu.2022.02.041

Gupta T, Gkiousias V, Bhutta MF. (2021). A systematic review of outcomes of remote consultation in ENT. Clinical Otolaryngology. 46(4): 699–719. doi: https://doi.org/10.1111/coa.13768

Guyatt GH, Oxman AD, Vist GE, et al. (2008). GRADE: an emerging consensus on rating quality of evidence and strength of recommendations. BMJ. 336(7650): 924. doi: 10.1136/bmj.39489.470347.AD

Henry TW, Maheu A, Sodha S, et al. (2022). Do postoperative telehealth visits require a high rate of redundant in-person evaluation after upper extremity surgery? Cureus. 14(1): e21462. doi: https://doi.org/10.7759/cureus.21462

Hutchings R. (2020). The impact of Covid-19 on the use of digital technology in the NHS. Nuffield Trust. Available at: https://www.nuffieldtrust.org.uk/files/2020-08/the-impact-of-covid-19-on-the-use-of-digital-technology-in-the-nhs-web-2.pdf.

Irarrázaval MJ, Inzunza M, Muñoz R, et al. (2021). Telemedicine for postoperative follow-up, virtual surgical clinics during COVID-19 pandemic. Surgical Endoscopy. 35(11): 6300–6. doi: https://doi.org/10.1007/s00464-020-08130-1

Jenkins JM, Halai M. (2021). CORR synthesis: What evidence is available for the continued use of telemedicine in orthopaedic surgery in the post-COVID-19 Era? Clinical Orthopaedics and Related Research. 479(4): 747–54. doi: https://doi.org/10.1097/corr.0000000000001444

Kolcun JPG, Ryu WHA, Traynelis VC. (2020). Systematic review of telemedicine in spine surgery. Journal of Neurosurgery. 1-10. doi: https://doi.org/10.3171/2020.6.Spine20863

Lightsey HMI, Crawford AM, Xiong GX, et al. (2021). Surgical plans generated from telemedicine visits are rarely changed after in-person evaluation in spine patients. The Spine Journal 21(3): 359–65. doi: https://doi.org/10.1016/j.spinee.2020.11.009

McMaster T, Wright T, Mori K, et al. (2021). Current and future use of telemedicine in surgical clinics during and beyond COVID-19: a narrative review. Annals of Medicine and Surgery. 66: 102378. doi: https://doi.org/10.1016/j.amsu.2021.102378

Moisan P, Barimani B, Antoniou J. (2021). Orthopedic surgery and telemedicine in times of COVID-19 and beyond: a review. Current Reviews in Musculoskeletal Medicine. 14(2): 155–9. doi: https://doi.org/10.1007/s12178-021-09693-9

Moola S, Munn Z, Tufanaru C, et al. (2020). Chapter 7: Systematic reviews of etiology and risk. In: Aromataris E & Munn Z (eds.) JBI Manual for Evidence Synthesis. JBI.

Natale J, Pascoe J, Horn C, et al. (2022). Teleconsultation versus traditional clinical assessment of patients undergoing circumcision: A retrospective cohort study. Journal of Clinical Urology. doi: https://doi.org/10.1177%2F20514158221088680

Nuffield Trust. (2022). NHS performance summary: February-March 2022. London: Nuffield Trust. Available at: https://www.nuffieldtrust.org.uk/news-item/nhs-performance-summary-february-march-2022 [Accessed 28th April 2022].

Page MJ, McKenzie JE, Bossuyt PM, et al. (2021). The PRISMA 2020 statement: an updated guideline for reporting systematic reviews. BMJ. 372: n71. doi: https://doi.org/10.1136/bmj.n71

Petersen W, Karpinski K, Backhaus L, et al. (2021). A systematic review about telemedicine in orthopedics. Archives of Orthopaedic and Trauma Surgery. 141(10): 1731–9. doi: https://doi.org/10.1007/s00402-021-03788-1

QualityWatch. (2020). The remote care revolution during Covid-19. London: The Health Foundation, Nuffield Trust,. Available at: https://www.nuffieldtrust.org.uk/files/2020-12/QWAS/digital-and-remote-care-in-covid-19.html#1 [Accessed 14th July 2022].

Raad M, Ndlovu S, Neen D. (2021). Assessment of the efficacy of telephone medicine consultations in trauma and orthopaedics during COVID-19 using the Ashford clinic letter score. Cureus. 13(3): e13871. doi: https://doi.org/10.7759/cureus.13871

Ryan R, Hill S. (2016). How to GRADE the quality of the evidence. Cochrane Consumers and Communication Group. Available at: https://colorectal.cochrane.org/sites/colorectal.cochrane.org/files/public/uploads/how_to_grade.pdf [Accessed 30th June 2022].

Scottish Intercollegiate Guidelines Network. (2019). Methodology checklist 3: cohort studies. Edinburgh: Healthcare Improvement Scotland. Available at: https://www.sign.ac.uk/what-we-do/methodology/checklists/ [Accessed 20th June 2022].

Sibanda V, Onubogu I, Raad M, et al. (2021). How effective are telephone and video consultations in shoulder and elbow clinics? analysis using an objective scoring tool. Cureus. 13(8): e17380. doi: https://doi.org/10.7759/cureus.17380

Smith SM, Jacobsen JHW, Atlas AP, et al. (2021). Telehealth in surgery: an umbrella review. ANZ Journal of Surgery. 91(11): 2360–75. doi: https://doi.org/10.1111/ans.17217

Technology Enabled Care Cymru. (2021). NHS Wales video consultation service - Phase 2a evaluation. Available at: https://digitalhealth.wales/news/phase-2a-evaluation.

Thomas J, O’Mara-Eves A, Harden A, et al. (2017). Chapter 8: Synthesis methods for combining and configuring textual or mixed methods data. An introduction to systematic reviews. London: Sage Publications Limited.

Tufanaru C, Munn Z, Aromataris E, et al. (2020). Chapter 3: Systematic reviews of effectiveness. In: Aromataris E MZ (ed.) JBI Manual for Evidence Synthesis. JBI.

Tully L, Case L, Arthurs N, et al. (2021). Barriers and facilitators for implementing paediatric telemedicine: rapid review of user perspectives. Frontiers in pediatrics. 9: 630365. doi: 10.3389/fped.2021.630365

Uppal A, Kothari AN, Scally CP, et al. (2022). Adoption of telemedicine for postoperative follow-up after inpatient cancer-related surgery. JCO oncology practice. March 9 (Online ahead of print): OP2100819. doi: https://doi.org/10.1200/op.21.00819

Virani S, Eastwood S, Holmes N, et al. (2021). Objective assessment of the efficacy of telephone medicine consultations in dispensing elective orthopaedic care using a novel scoring tool. The Surgeon. 19(5): e175–e82. doi: https://doi.org/10.1016/j.surge.2020.09.008

Welsh Government. (2022). NHS activity and performance summary: February and March 2022. Cardiff Welsh Government. Available at: https://gov.wales/nhs-activity-and-performance-summary-february-and-march-2022-html [Accessed 28th April 2022].

Ye IB, Thomson AE, Donahue J, et al. (2022). Similar accuracy of surgical plans after initial in-person and telemedicine evaluation of spine patients. World neurosurgery. May 27 (online ahead of print): S1878-8750(22)00731-8. doi: https://doi.org/10.1016/j.wneu.2022.05.091

